# The TARCiS statement: Guidance on terminology, application, and reporting of citation searching

**DOI:** 10.1101/2023.10.25.23297543

**Authors:** Julian Hirt, Thomas Nordhausen, Thomas Fuerst, Hannah Ewald, TARCiS study group, Christian Appenzeller-Herzog

**Affiliations:** Pragmatic Evidence Lab, Research Center for Clinical Neuroimmunology and Neuroscience Basel (RC2NB), University Hospital Basel and University of Basel, Basel, Switzerland; Department of Health, Eastern Switzerland University of Applied Sciences, St. Gallen, Switzerland; Department of Clinical Research, University Hospital Basel and University of Basel, Basel, Switzerland; Institute of Health and Nursing Science, Medical Faculty, Martin Luther University Halle-Wittenberg, Halle (Saale), Germany; University Medical Library, University of Basel, Basel, Switzerland; TARCiS study group: Alison Avenell (University of Aberdeen, United Kingdom), Alison Bethel (University of Exeter, United Kingdom), Andrew Booth (University of Sheffield and University of Limerick, United Kingdom), Christopher Carroll (University of Sheffield, United Kingdom), Justin Clark (Bond University, Australia), Julie Glanville (Glanville.info, United Kingdom), Su Golder (University of York, United Kingdom), Elke Hausner (Institute for Quality and Efficiency in Health Care (IQWiG), Germany), Tanya Horsley (Royal College of Physicians and Surgeons, Canada), David Kaunelis (Canadian Agency for Drugs and Technologies in Health (CADTH), Canada), Shona Kirtley (University of Oxford, United Kingdom), Irma Klerings (Donau University, Austria), Jonathan Koffel (United States), Paul Levay (National Institute for Health and Care Excellence (NICE), United Kingdom), Kathrine McCain (Drexel University, United States), Maria-Inti Metzendorf (Heinrich-Heine University Duesseldorf, Germany), David Moher (University of Ottawa, Canada), Linda Murphy (University of California at Irvine, United States), Melissa Rethlefsen (University of New Mexico, United States), Amy Riegelman (University of Minnesota, United States), Morwenna Rogers (University of Exeter, United Kingdom), Margaret Sampson (Children’s Hospital of Eastern Ontario, Canada), Jodi Schneider (University of Illinois at Urbana-Champaign, United States), Terena Solomons (Curtin University, Australia), Alison Weightman (Cardiff University, United Kingdom)

**Keywords:** Methods[MeSH], Research Design[MeSH], Systematic Review as Topic[MeSH], Delphi Technique[MeSH], Information Storage and Retrieval[MeSH], Citation Searching, Supplementary Search Methods

## Abstract

Evidence syntheses adhering to systematic literature searching techniques are a cornerstone of evidence-based health care. Beyond term-based searching in electronic databases, citation searching is a prevalent search technique to identify relevant sources of evidence. However, for decades, citation searching methodology and terminology has not been standardized. We performed an evidence-guided four-round Delphi consensus study with 27 international methodological experts in order to develop the Terminology, Application, and Reporting of Citation Searching (TARCiS) statement. TARCiS comprises ten specific recommendations on when and how to conduct and report citation searching in the context of systematic literature searches and four research priorities. We encourage systematic reviewers and information specialists to incorporate TARCiS into their standardized workflows.

## INTRODUCTION

Synthesizing scientific evidence by pursuing citation relationships of references (citation searching) was the underlying objective when the Science Citation Index, the antecedent of today’s Web of Science, was introduced in 1963.^1^ Although the availability of electronic citation indexes has increased, evidence syntheses in systematic reviews do not primarily rely on citation searching for literature retrieval but rather on text- and keyword-based search methods.^2^ When used in systematic review workflows, citation searching traditionally constitutes a supplementary search technique that builds on an initial set of references from the primary database search (seed references).^3^

Citation searching is an umbrella term that entails various methods of citation-based literature retrieval (Figure 1). Checking references cited by the seed references, also known as backward citation searching, is the most prevalent and a mandatory step in Cochrane reviews.^4^ In opposite direction (forward citation searching), systematic reviewers can also assess the eligibility of articles that cite the seed references. Backward and forward citation searching are known as direct citation searching (Figure 1). They can be supplemented by indirect retrieval methods, namely by co-citing citation searching (retrieving articles that share cited references with a seed reference) and co-cited citation searching (retrieving articles that share citing references with a seed reference). Figure 1 illustrates these terms.

**Figure 1.**
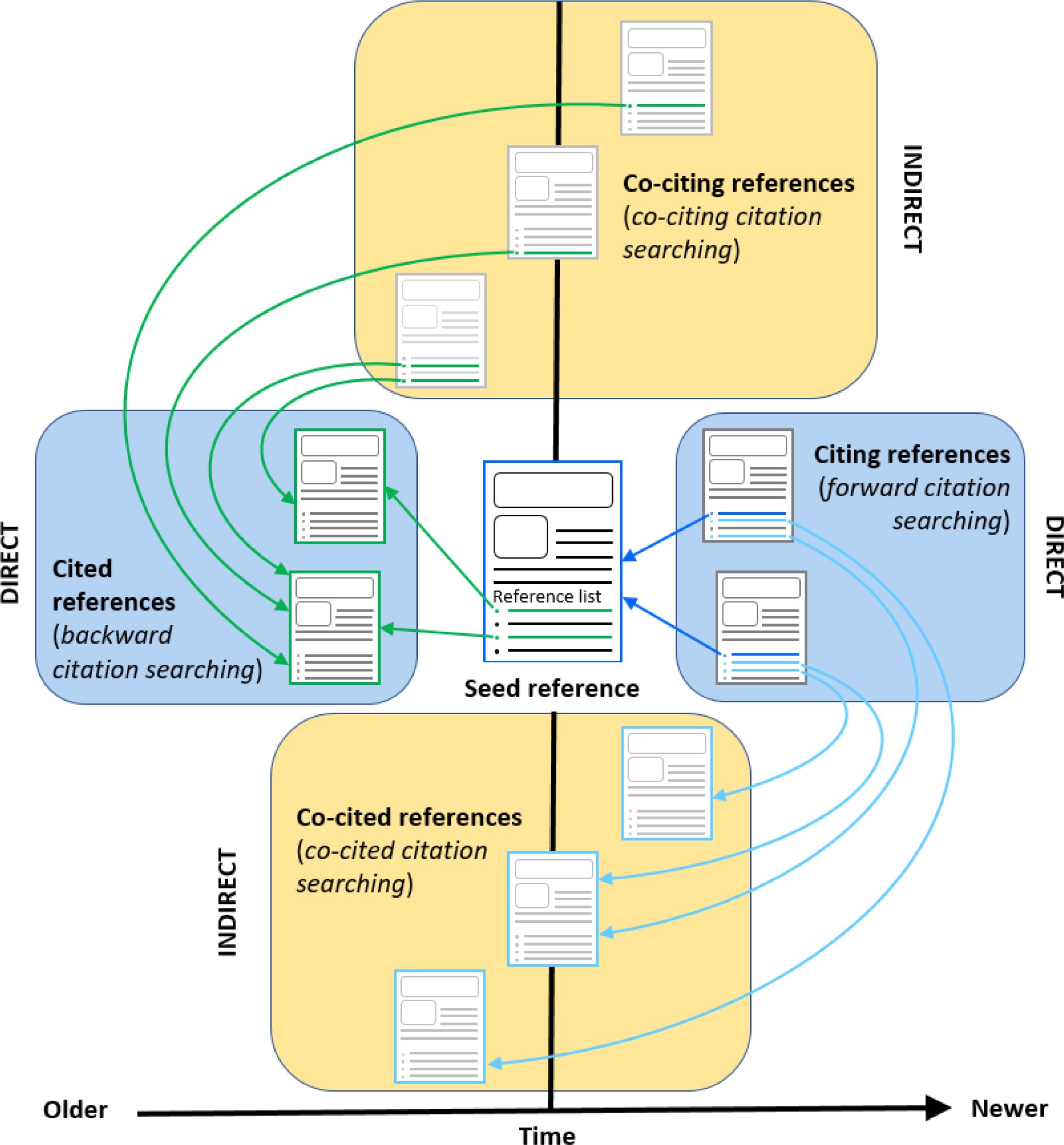
Overview of citation searching methods

The conduct of systematic reviews is prominently guided by standard recommendations such as those in the Cochrane handbook,^4^ whereas their reporting is standardized by the Preferred Reporting Items for Systematic reviews and Meta-Analyses (PRISMA) statement.^5^ In stark contrast and despite its application by systematic reviewers for decades, standardized methodology and terminology for citation searching is not available. Only for the latter of the following three aspects (i) “when to do citation searching”, (ii) “how to conduct citation searching”, and (iii) “how to report citation searching”, limited guidance exists by the PRISMA for Searching (PRISMA-S) extension.^6^ Unsurprisingly, methodological studies display marked heterogeneity in terms of citation searching terminology and recommended best practices,^7^ thus impeding evidence-based decision-making on when and how to best incorporate citation searching into researchers’ systematic review workflows.

We systematically collected evidence on the use, benefit, and reporting of citation searching^7^ and subjected it to a four-round online Delphi study. Together with the Terminology, Application, and Reporting of Citation Searching (TARCiS) study group, an international panel of methodological experts, we aimed to develop consensus for recommendations on (i) when and (ii) how to conduct citation searching and (iii) how to report it, including a consensus set of citation searching terminology. We framed research priorities for future methodological development of citation searching in the context of systematic literature searches.

## METHODS

To develop the TARCiS statement, our core team of information specialists and researchers followed a stepwise approach comprising a systematic synthesis of methodological literature (step 1; reported in detail in a separate publication)^7^ and a Delphi study (step 2; reported in this publication). We prespecified our methods.^8^ ^9^ A study flow diagram is shown in Figure 2.

**Figure 2.**
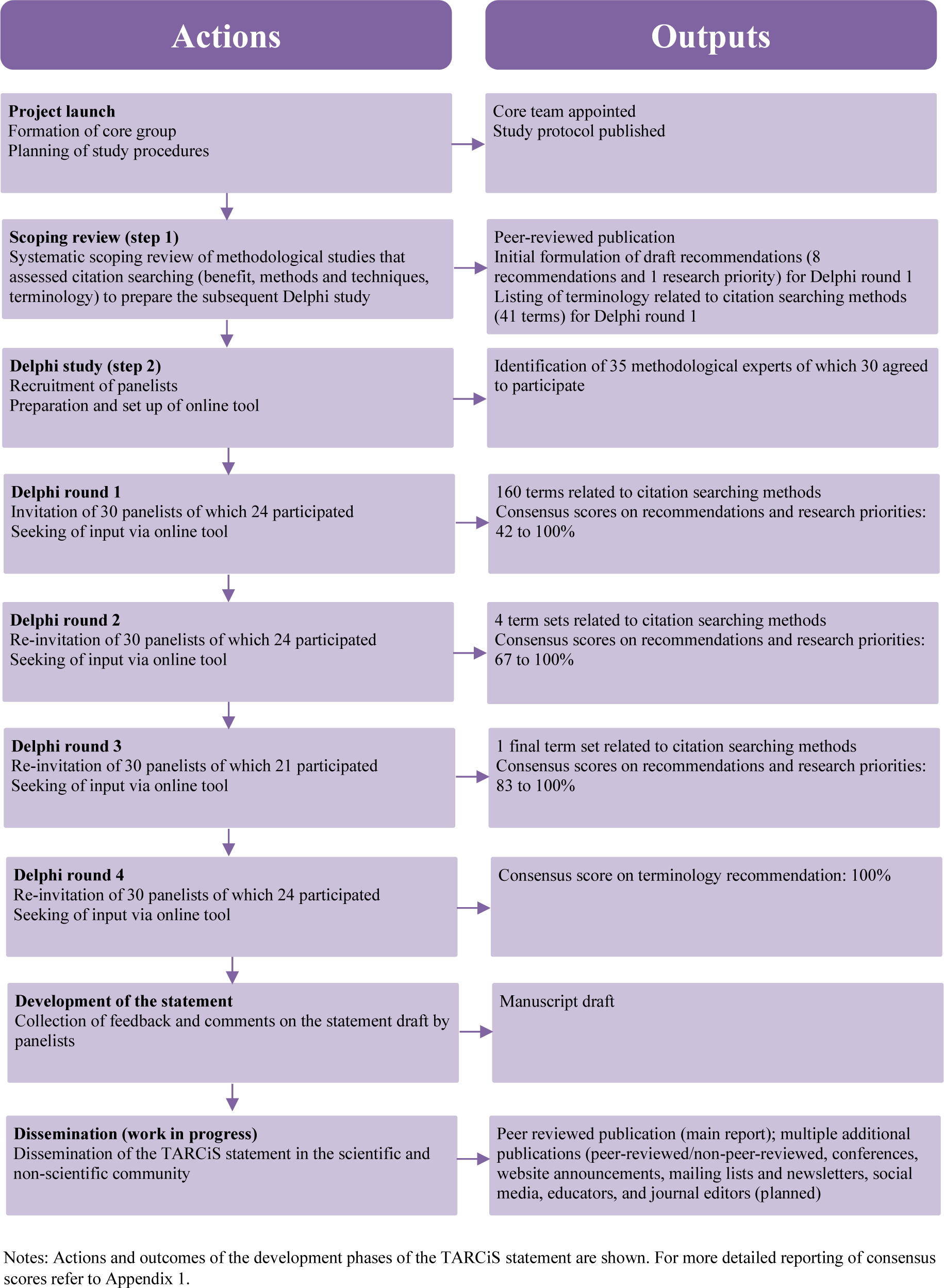
Flow diagram of the development process of the TARCiS statement

### Step 1: Scoping review

We conducted a scoping review following the framework outlined by Arksey and O’Malley^10^ on the terminology that describes citation searching, the methods and tools used for citation searching, and its benefit. We considered methodological studies of any design that aimed to assess the role of citation searching and/or compared multiple citation searching methods (e.g., backward vs. forward citation searching) and/or compared technical uses of citation searching (e.g., forward citation searching using Scopus vs. Google Scholar) within health-related topics. We searched five bibliographic databases, conducted backward and forward citation searches of eligible studies and pertinent reviews, and consulted librarians and information specialists for further eligible studies. The results were summarized by descriptive statistics and narratively.

### Step 2: Delphi study

To develop consensus on recommendations and research priorities as tentatively derived from the results of Step 1,^7^ we performed a multi-stage online Delphi study. Delphi refers to a structured process where collective knowledge from an expert panel is synthesized using a series of questionnaires, each one adapted based on the responses to a previous version.^11–13^ We recruited an international panel of individuals experienced in conducting and/or reporting citation searching methods. For this, we invited authors of methodological studies^7^ and methodological experts from international systematic review organizations or from our professional networks by e-mail to participate in the Delphi study.

The Delphi study comprised four prespecified rounds.^9^ The first round was pretested by four non-study-related academic affiliates. Each round covered four to five thematic parts (Appendix 2; see also Table 1 parts A – E). Briefly, part A dealt with the terminology framework to describe citation searching methods in eight domains (for details, refer to Table 4 in Hirt et al.^7^). Part B contained pre-formulated recommendations on conduct and reporting of citation searching. Each recommendation was supported by a rationale and explanation text that were also subjected to collective consensus finding. Part C addressed research priorities that were also derived from the scoping review.^7^ Part D contained a free text field to collect general comments to the core team. Part E was designed to collect sociodemographic information and was limited to Delphi round 1.

**Table 1.** Data collection through the Delphi rounds.

| Part | Round 1 | Round 2 | Round 3 | Round 4 |
| --- | --- | --- | --- | --- |
| **A: Terminology framework (8 terminology domains <sup>+</sup>)** | Terminology suggestions informed by scoping review<sup>7</sup> per domain<br>Free text field for additional suggestions per domain | Selection of preferred terms per domain (minimum 1 term, maximum 3 terms) | Selection of a preferred term set | N/A<br>[the term set with the most votes as resulted in Round 3 was incorporated into TARCiS recommendation 1 and agreement was rated as for other recommendations] |
| **B: Recommendations, each accompanied by a rationale and explanation** | Agreement rating for all recommendations *<br>Comment to support rating<br>Comment on rationale and explanation<br>Free text field for additional recommendations | Agreement rating for remaining amended recommendations *<br>Agreement rating for additional recommendations <sup>§</sup><br>Comment to support rating<br>Comment on rationale and explanation | Agreement rating for remaining amended recommendations *<br>Comment to support rating<br>Comment on rationale and explanation | Agreement rating for remaining amended rationales and explanations *<br>Comment on rationale and explanation<br>Free text field for referencing suggestions |
| **C: Research priorities, accompanied by a rationale and explanation where necessary** | Agreement rating for all research priorities *<br>Comment to support rating<br>Comment on rationale and explanation<br>Free text field for additional research priorities | Agreement rating for remaining amended research priorities *<br>Comment to support rating | N/A | Free text field for referencing suggestions |
| **D: Other comments** | Free text field for additional comments | Free text field for additional comments | Free text field for additional comments | N/A |
| **E: Sociodemographic information** | Open-ended questions<br>Single-choice questions | N/A ** | N/A ** | N/A ** |

\* Strongly agree, agree, disagree, strongly disagree; \*\* Of participants who missed Delphi round 1, we collected sociodemographic information via e-mail.
<sup>+</sup> As derived from Figure 1 using neutral terminology: umbrella term; sub-method retrieving and screening cited references; sub-method retrieving and screening cited references by manually reviewing reference lists; sub-method retrieving and screening citing references; sub-method retrieving and screening co-cited references; sub-method retrieving and screening co-citing references; iterative repetition of a citation-based method; relevant articles known beforehand.
<sup>§</sup> None of the additional recommendations attained accordance for inclusion into the TARCiS statement.
Abbreviations: N/A = Not applicable.

We distributed the Delphi survey via Unipark/EFS survey^14^ using system-generated e-mails that contained a personalized survey link. This link allowed the core team to individually remind panelists to participate. The duration of each round was three weeks. Non-participating panelists were recorded as non-participators for a given round. Panelists who missed all rounds were recorded as non-responders. The survey entailed a variety of data collection techniques (agreement rating, free text, single-choice question, open-ended question, comment). We considered panelists had reached consensus when at least 75% agreed or strongly agreed. Recommendations and research priorities that had not yet reached consensus were refined for the subsequent Delphi round. These adaptations were based on the panelists’ comments. In rare cases, when additional valid panelists’ suggestions for reformulation of rationale and explanation texts were submitted, recommendations that already reached the agreement threshold were also adapted and forwarded to the next Delphi round.

We used descriptive statistics for numeric data (or for data that could be converted into numbers) and narratively summarized the results using numbers and/or percentages.

We did not anticipate panelists to be vulnerable and, with regard to the Swiss Human Research Act, our research did not concern human diseases nor the structure and function of the human body.^15^ Thus, we did not apply for ethical approval. The landing page of each Delphi round contained information on the study aim, data management, and data security. Panelists’ assessments were anonymous to other panelists but open to the study team. Panelists were aware that taking part indicated consent to participate. They did not receive an incentive for participation and could leave the process at any time.

### Deviations from the Delphi study protocol

To ensure consistent terminology throughout the guidance, we decided to present the three terms that received the most votes in Delphi round 2 as four consistent term sets. Second, instead of using SosciSurvey^16^ as a survey tool,^8^ we switched to Unipark/EFS survey,^14^ which provided enhanced design and functional features. Third, in addition to personalized e-mails (person-based approach), we originally intended to recruit panelists using professional mailing lists and central requests to systematic review organizations (organization-based approach).^8^ However, as we had already recruited sufficient panelists using the person-based approach (including individuals that were affiliated with systematic review organizations), we waived the organization-based approach.

### Patient and public involvement

We did not involve patients nor members of the public in formulating the research objectives, designing the study, interpreting the results, or writing the manuscript.

## RESULTS

### Step 1: Scoping review

We identified 47 methodological studies that assessed the use, benefit, and reporting of citation searching. In 45 studies (96%), the use of citation searching showed an added value. Thirty-two studies (68%) analyzed the impact of citation searching in one or more previous systematic reviews. Application, terminology, and reporting of citation searching were heterogeneous. Details on the results of the scoping review can be found elsewhere.^7^

### Step 2: Delphi study

#### Recruitment and characteristics of panelists

We identified and contacted 35 experts, 30 of whom declared interest in participating and were in turn invited to Delphi round 1. Three of the 30 panelists (10%) did not take part in any Delphi round and were counted as non-responders. The demographic and professional characteristics of the 27 responding panelists are summarized in Table 2.

**Table 2.**
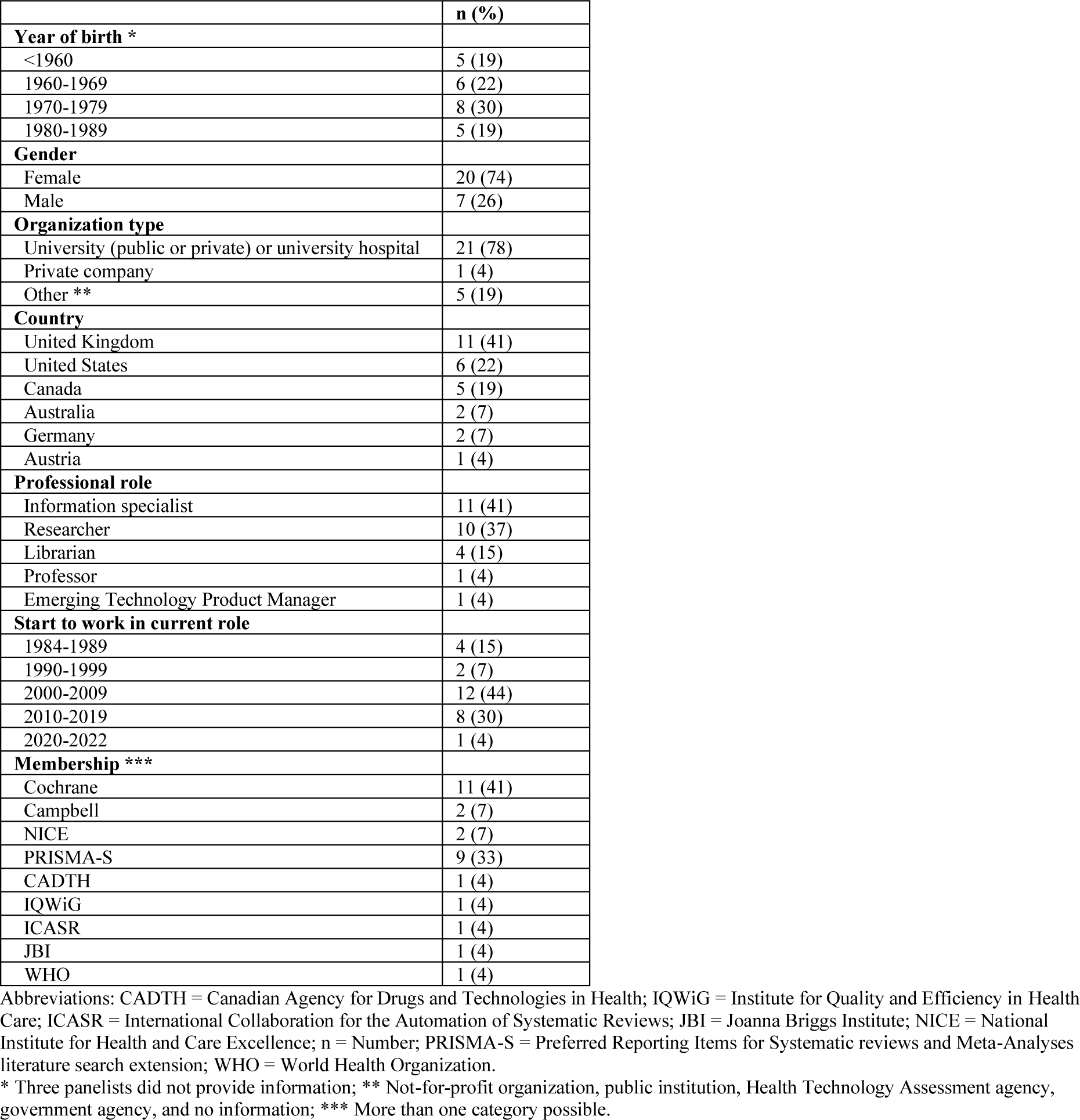
Characteristics of the 27 panelists.

#### The TARCiS statement

Items for data collection through the four Delphi rounds in parts A – E are summarized in Table 1. Informed by the scoping review^7^ and our expertise, the Delphi study started with 41 terminology framework terms, eight draft recommendations with rationale texts on the conduct and reporting of citation searching, and one research priority. After Delphi round 4, the finalized TARCiS statement comprised ten recommendations with rationale and explanation texts and four research priorities that reached consensus scores between 83% and 100%. See Figure 2 and Appendix 1 for details on content and consensus scores in rounds 1 to 4. An overview of all fourteen TARCiS items omitting rationale and explanation texts is presented in Table 3. A reporting item checklist based on TARCiS recommendation 10 is available in Appendix 3 and on Open Science Framework.^17^

**Table 3.**
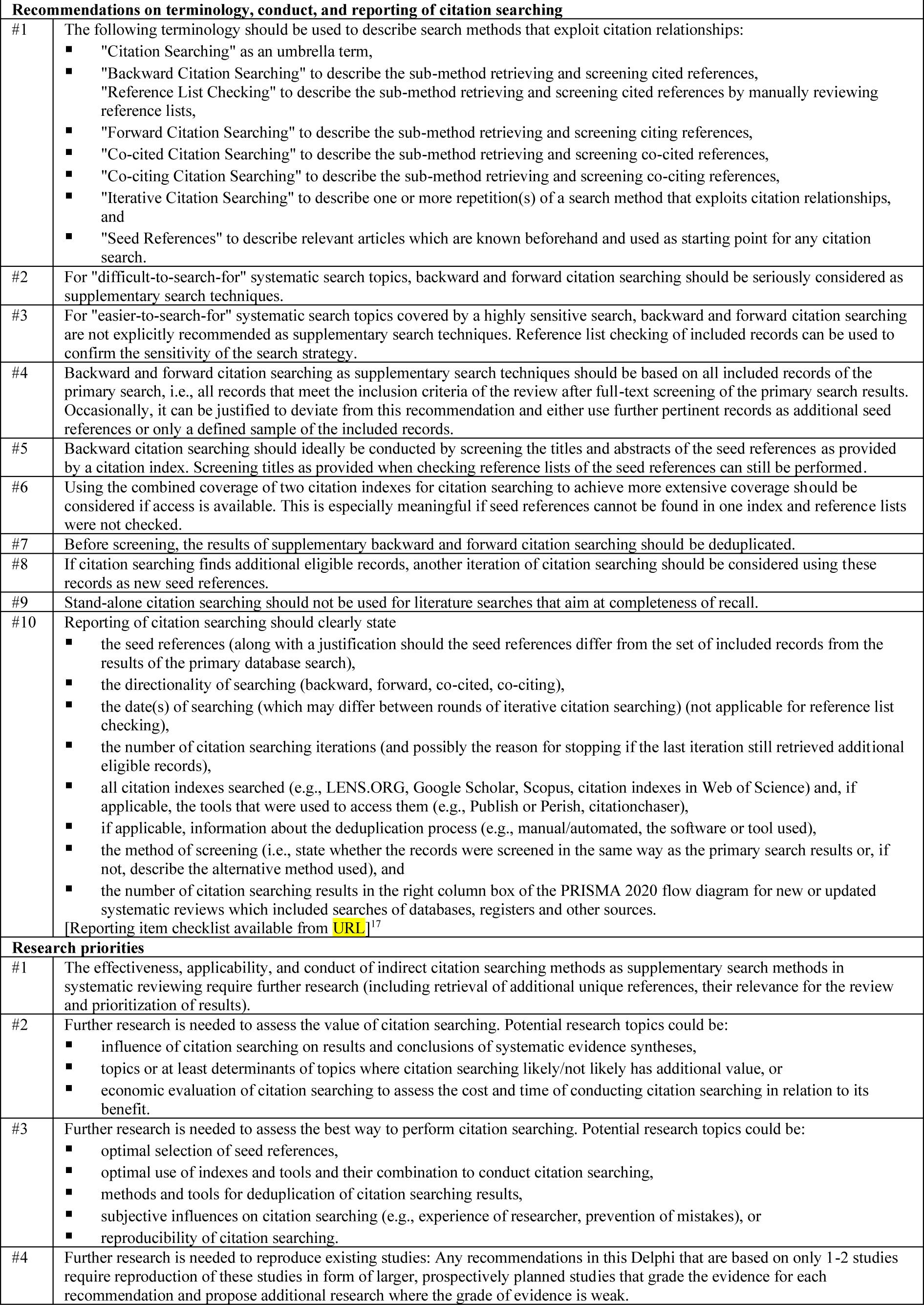
TARCiS statement.

*Final recommendations, rationale and explanations, and research priorities*

##### Recommendation 1

The following terminology should be used to describe search methods that exploit citation relationships:

- *”Citation Searching”* as an umbrella term,
- *”Backward Citation Searching”* to describe the sub-method retrieving and screening cited references,

*”Reference List Checking”* to describe the sub-method retrieving and screening cited references by manually reviewing reference lists,

- *”Forward Citation Searching”* to describe the sub-method retrieving and screening citing references,
- *”Co-cited Citation Searching”* to describe the sub-method retrieving and screening co-cited references,
- *”Co-citing Citation Searching”* to describe the sub-method retrieving and screening co-citing references,
- *”Iterative Citation Searching”* to describe one or more repetition(s) of a search method that exploits citation relationships, and
- *”Seed References”* to describe relevant articles which are known beforehand and used as starting point for any citation search.

##### Rationale and explanation supporting recommendation 1

As compiled in a recent scoping review,^7^ the reporting of citation searching methods is frequently unclear and far from being standardized. For example, “citation searching”, “snowballing”, or “co-citation searching” are sometimes used as methodological umbrella terms but also to denote a specific method such as backward or forward citation searching.^7^ For clarity, standardized vocabulary is needed. The set of terms brought forward in this recommendation is consistent in itself as well as with the terminology used in PRISMA-S and PRISMA 2020 guidelines^6^ ^18^ and hence well suited for uniform reporting of citation searching.

##### Recommendation 2

For “difficult-to-search-for” systematic search topics, backward and forward citation searching should be seriously considered as supplementary search techniques.

##### Rationale and explanation supporting recommendation 2

Evidence indicates that the ability of citation searching as a supplementary search technique to find additional unique records in a systematic literature search varies with the topic.^7^ Searches for particular study designs (qualitative, mixed-method, observational, prognostic, or diagnostic test studies) or health science topics such as non-pharmacological, non-clinical, public health, policy making, service delivery, or alternative medicine have been linked with effective supplementary citation searching.^19–22^ The underlying reasons are manifold and include poor transferability of the topic to text-based searching (e.g., owing to poor conceptual clarity, inconsistent terminology, or vocabulary overlaps with other topics).^23^ The ability of citation searching to find any publication type including unpublished or grey literature or literature that is not indexed in major databases (e.g., concerning a developing country) may also be relevant.^24^ However, a clear categorization of “difficult-to-search-for” topics is currently not possible and it remains for the review authors themselves to judge whether their review topic is likely to fall into this category. We recommend that persons conducting the search who have difficulty assessing whether the topic is difficult-or easier-to-search-for always opt for citation searching or consult an experienced information specialist.^25^ If for whatever reason the search strategy does not exhaustively capture the topic, backward and forward citation searching may compensate for the potential loss of information to some extent.

##### Recommendation 3

For “easier-to-search-for” systematic search topics covered by a highly sensitive search, backward and forward citation searching are not explicitly recommended as supplementary search techniques. Reference list checking of included records can be used to confirm the sensitivity of the search strategy.

##### Rationale and explanation supporting recommendation 3

Evidence indicates that the ability of citation searching as a supplementary search technique to find additional unique references in a systematic literature search varies with the search topic.^7^ Searches for clearly defined clinical interventions as part of Participant-Intervention-Comparison-Outcome (PICO)-questions have been linked with less effective supplementary citation searching, especially when the search strategies are sensitive and conducted in several databases. However, a clear categorization of “easier-to-search-for” topics is currently not possible and it remains for the review authors themselves to judge whether their review topic is likely to fall into this category.

By checking reference lists within the full-texts of seed references, review authors can test the sensitivity of their primary search strategy (i.e., electronic database search).^26^ Should no additional relevant, unique studies be found, the primary search may have been sensitive enough. Should additional relevant, unique studies be found, it may be an indication that the primary search was not sensitive enough.

We recommend that persons conducting the search who have difficulty assessing whether the topic is difficult-or easier-to-search-for opt for citation searching or consult an experienced information specialist.^25^ If for whatever reason the search strategy does not exhaustively capture the topic, backward and forward citation searching may compensate for the potential loss of information to some extent.

##### Recommendation 4

Backward and forward citation searching as supplementary search techniques should be based on all included records of the primary search, i.e., all records that meet the inclusion criteria of the review after full-text screening of the primary search results. Occasionally, it can be justified to deviate from this recommendation and either use further pertinent records as additional seed references or only a defined sample of the included records.

##### Rationale and explanation supporting recommendation 4

The more seed references are used, the better are the chances that citation searching finds additional relevant unique records. While using only a sample of the included records as seed references may be enough, there is currently no evidence that could help decide how many seeds are needed or how to decide which may perform better. Hence, we recommend using all the records that meet the inclusion criteria of the review after full-text screening of the primary database search results.

However, review authors may deviate from this recommendation if they deal with a very small or large number of included records. A very small number of included records may not yield additional relevant records or only have limited value. In this case, review authors could use further records as seed references for citation searching (e.g., systematic reviews on the topic that were flagged during the screening phase).^27^ A very large number of included records could lead to too many records to screen. In this case, review authors may use a selected sample of included records as seed references for citation searching. In the event of such deviation, authors should describe their rationale and sampling method (e.g., random sample).

##### Recommendation 5

Backward citation searching should ideally be conducted by screening the titles and abstracts of the seed references as provided by a citation index. Screening titles as provided when checking reference lists of the seed references can still be performed.

##### Rationale and explanation supporting recommendation 5

Citation searching workflows encompass two consecutive steps: retrieval of records and screening of retrieved records for eligibility. When using an electronic citation index for citation searching, retrieval and screening are usually separated. While forward citation searching requires a citation index, backward citation searching can also be performed by manually checking the reference lists of the seed references. Reference list checking is sometimes part of an established workflow, e.g., done during eligibility assessment of the full-text record or during data extraction.^26^ Merging these two steps has the benefit that researchers know the context in which a reference was used and that all references can be screened. However, reference list checking has three disadvantages: (i) the retrieval and screening phases are no longer separated which makes reporting of the methods/results difficult and unclear, (ii) citations from reference list checking cannot be deduplicated against each other and/or against the primary search results which may add an unnecessarily high workload (see recommendation 7), and (iii) eligibility assessments are restricted to the titles (instead of titles and abstracts) which could lead to relevant records being overlooked due to uninformative titles mentioned in vague contexts.

In recent years, online citation searching options via citation indexes or free to access citation searching tools have become more readily available leading to faster and easier procedures.^28–31^ More and even better tools to facilitate this workflow are expected in the future. Combining citation searching via citation indexes with automated deduplication (free online tools available)^32–34^ makes this recommendation feasible. A caveat is that a search in a single citation index will in most cases fail to retrieve all the cited references.^35^ ^36^ Thus, references to some documents (such as websites, registry entries or grey literature) that are less likely to be indexed in databases may only be retrievable by checking reference lists or only in some citation indexes.^3^

##### Recommendation 6

Using the combined coverage of two citation indexes for citation searching to achieve more extensive coverage should be considered if access is available. This is especially meaningful if seed references cannot be found in one index and reference lists were not checked.

##### Rationale and explanation supporting recommendation 6

A single citation index or citation analysis tool may not cover all seed references and very likely will not find all the citing and cited literature. The reasons for this are that citation indexes do not offer 100% coverage as some references are currently not indexed in one or several citation index(es)^37^ as well as data quality issues.^38^ Evidence indicates that when using more than one citation index for citation searching, the results of the different indexes can complement each other.^39–41^ Thus, retrieval of backward and forward citation searching results from more than one citation index or citation analysis tool (e.g., TheLens via citationchaser, Scopus, citation indexes in Web of Science) followed by deduplication (see recommendation 6) can increase the sensitivity of citation searching. It is similar to the complementary effect of using multiple electronic databases for the primary database search, which is the gold-standard in systematic search workflows.^4^ In recent years, online citation searching options have increased and many open access tools make rapid electronic citation searching universally accessible.^28–31^

##### Recommendation 7

Before screening, the results of supplementary backward and forward citation searching should be deduplicated.

##### Rationale and explanation supporting recommendation 7

The concept of citation searching as a supplementary search method relies on the notion that reference and cited-by lists of eligible references are topically related to these references.^7^ This implies a considerable degree of overlap within these lists leading to several duplicates. Furthermore, the overlap likely also extends to the results of the primary database search that was performed on the same topic. Based on these considerations and on the fact that the results of the primary database search have already been screened for eligibility, the screening load of citation searching results can be significantly cut by removing those references that have already been screened for eligibility (deduplication against the primary database search) and those that appear as duplicates during citation searching.^35^ Depending on the method of deduplication, this can be done in one go.

While deduplication can be conducted manually, nowadays standard bibliographic management software and specialized tools provide automated deduplication solutions, allowing for easier and faster processing.^35^ ^42^ ^43^

If citation searching leads to only a very small number of results, omission of the deduplication step can be considered to save time and administrative effort.

##### Recommendation 8

If citation searching finds additional eligible records, another iteration of citation searching should be considered using these records as new seed references.

##### Rationale and explanation supporting recommendation 8

Citation searching methods can be conducted over one or more iteration(s), a process we refer to as iterative citation searching.^44^ The first iteration is based on the original seed references (see recommendation 4). If eligibility screening of the results of this first iteration leads to the inclusion of further eligible records, these records serve as new seed references for the second iteration and so forth. There is evidence that conducting iterative citation searching can contribute to the identification of more eligible records.^7^ ^44–46^

Since iterations beyond the first round of citation searching require additional time and effort and may interrupt the ongoing review process, the decision in favor of or against further iterations should be guided by an informal cost-benefit assessment. Relevant factors to be assessed include the review topic (difficult-or easier-to-search-for), sensitivity of the primary search, aim for completeness of the literature search, and the estimated potential benefit of the iteration(s) (e.g., based on the number/percentage of included records found with the previous citation searching iteration).

Review authors should report the number of iterations and possibly the reason for stopping if the last iteration still retrieved additional eligible records.

Please note that stating *”citation searching was done on all included records”* can lead to confusion. Most authors may mean all records included after full-text screening of the primary search results. However, strictly speaking, *”all included records”* also includes the records retrieved via citation searching. The latter interpretation implies that iterative citation searching is required until the last iteration leads to no further identification of eligible records.

As outlined in the rationale of recommendation 7, results of citation searching iterations can be deduplicated against all previously retrieved records to reduce the screening load.

##### Recommendation 9

Stand-alone citation searching should not be used for literature searches that aim at completeness of recall.

##### Rationale and explanation supporting recommendation 9

We refer to stand-alone citation searching when any form of citation searching is used as the primary search method without extensive prior database searching.^7^ This is contrary to citation searching as a supplementary search method to a primary database search. Seed references for stand-alone citation searching could, for example, be records from researchers’ personal collections or retrieved from less sensitive literature searches. Stand-alone citation searching can be based on a broad set of seed references. It can comprise backward and forward citation searching as well as indirect methods that collect co-citing and co-cited references.

When study authors have replicated published systematic reviews with stand-alone citation searching, they mostly missed literature that was included in the systematic review.^28^ ^47–49^ Since search methods for systematic reviews and scoping reviews should aim at completeness of recall, stand-alone citation searching is not a suitable method for these types of literature review.

##### Recommendation 10

Reporting of citation searching should clearly state

- the seed references (along with a justification should the seed references differ from the set of included records from the results of the primary database search),
- the directionality of searching (backward, forward, co-cited, co-citing),
- the date(s) of searching (which may differ between rounds of iterative citation searching) (not applicable for reference list checking),
- the number of citation searching iterations (and possibly the reason for stopping if the last iteration still retrieved additional eligible records),
- all citation indexes searched (e.g., LENS.ORG, Google Scholar, Scopus, citation indexes in Web of Science) and, if applicable, the tools that were used to access them (e.g., Publish or Perish, citationchaser),
- if applicable, information about the deduplication process (e.g., manual/automated, the software or tool used),
- the method of screening (i.e., state whether the records were screened in the same way as the primary search results or, if not, describe the alternative method used), and
- the number of citation searching results in the right column box of the PRISMA 2020 flow diagram for new or updated systematic reviews which included searches of databases, registers and other sources.

##### Rationale and explanation supporting recommendation 10

The relevant guidance for researchers conducting citation searching in systematic literature searching can be found in item 5 of PRISMA-S.^6^

Accordingly, required reporting items are the directionality of citation searching (examination of cited or citing references), methods and resources used for citation searching (bibliographies in full text articles or citation indexes), and the seed references that citation searching was performed upon.^6^ Additional information for the reporting of citation searching can be found in PRISMA-S items 1 (database name), 13 (dates of searches), and 16 (deduplication).^6^ While PRISMA-S can be seen as the minimum reporting standard for citation searching as a supplementary search technique, other important elements that emerged from our scoping review^7^ need to be reported to achieve full transparency and/or reproducibility. These elements are listed in recommendation 10 as a supplement to PRISMA-S to comprehensively guide the reporting of supplementary citation searching in systematic literature searching.

Concerning reporting of citation searching results in the PRISMA 2020 flow diagram,^50^ two variants are possible: (i) reporting of deduplicated records only which are additional to the primary search results or 1. (ii) reporting of all retrieved records followed by insertion of an additional box where the number of deduplicated records is reported.

Please note that the detail of the citation searching methods do not have to be reported in the main methods of a study. Detailed search information can be provided in an appendix or an online public data repository.

##### *Example 1* for good reporting

”As supplementary search methods, we performed […] direct forward and backward CT of included studies and pertinent review articles that were flagged during the screening of search results (on February 10, 2021). For forward CT, we used Scopus, Web of Science [core collection as provided by the University of Basel; Editions = SCI-EXPANDED, SSCI, A&HCI, CPCI-S, CPCI-SSH, BKCI-S, BKCI-SSH, ESCI, CCR-EXPANDED, IC], and Google Scholar. For *backward CT, we used Scopus and, if seed references were not indexed in Scopus, we manually extracted the seed references’ reference list. We iteratively repeated forward and backward CT on newly identified eligible references until no further eligible references or pertinent reviews could be identified (three iterations; the last iteration on May 5, 2021).”*^7^

##### Example 2 for good reporting

*”To supplement the database searches, we performed a forward (citing) and backwards (cited) citation analysis on 2 August 2022 using SpiderCite (*https://sr-accelerator.com/#/spidercite).”^51^

##### Example 3 for good reporting

*”Reference lists of any included studies and retrieved relevant SRs published in the last five years were checked for any eligible studies that might have been missed by the database searches.”*^52^

##### Research priority 1

The effectiveness, applicability, and conduct of indirect citation searching methods as supplementary search methods in systematic reviewing require further research (including retrieval of additional unique references, their relevance for the review and prioritization of results).

##### Rationale and explanation supporting research priority 1

Indirect citation searching involves the collection and screening for eligibility of records that share references in their bibliography or citations with one of the seed references (i.e., co-citing or co-cited references).^8^ Indirect citation searching typically retrieves a large volume of records to be screened.^47^ ^49^ Therefore, prioritization algorithms for the screening of records and cut-offs that may discriminate between potentially relevant and non-relevant records have been proposed that aim at reducing the workload of eligibility screening.^28^ ^48^ The methodological studies that have pioneered indirect citation searching methods for health-related topics have so far exclusively focused on stand-alone citation searching.^7^ It is currently unclear whether the added workload and resources for searching and screening warrant indirect citation searching methods as supplementary search techniques in systematic reviews of any type (qualitative/quantitative, difficult/easier-to-search-for).

##### Research priority 2

Further research is needed to assess the value of citation searching. Potential research topics could be:

- influence of citation searching on results and conclusions of systematic evidence syntheses,
- topics or at least determinants of topics where citation searching is likely/not likely to have additional value, or
- economic evaluation of citation searching to assess the cost and time of conducting citation searching in relation to its benefit.

##### Research priority 3

Further research is needed to assess the best way to perform citation searching. Potential research topics could be:

- optimal selection of seed references,
- optimal use of indexes and tools and their combination to conduct citation searching,
- methods and tools for deduplication of citation searching results,
- subjective influences on citation searching (e.g., experience of researcher, prevention of mistakes), or
- reproducibility of citation searching.

##### Research priority 4

Further research is needed to reproduce existing studies: Any recommendations in this Delphi that are based on only 1-2 studies require reproduction of these studies in the form of larger, prospectively planned studies that grade the evidence for each recommendation and propose additional research where the grade of evidence is weak.

## DISCUSSION

### TARCiS recommendations and research priorities

In keeping with our study aims, the TARCiS guideline covers three aspects of citation searching in the context of systematic literature searches. It recommends when to conduct citation searching, how to conduct citation searching, and how to report citation searching.

In systematic evidence syntheses, citation searching techniques can be used to fill gaps in the results of the primary database search, but their application is not universally indicated. TARCiS recommendations 2 and 3 provide critical assistance on whether or not a systematic search is likely to benefit from the use of citation searching. Systematic searchers of defined pharmaceutical interventions, for instance, may take from this guidance that they could skip citation searching as their primary database search may already allow for high sensitivity at reasonable specificity and expedite other supplementary search techniques such as clinical trial registry searching.^53^ This is because we do not recommend the use of citation searching in “easier-to-search-for” topics and as formulated in research priority 2, more research is needed to pinpoint and reliably discriminate “easier-to-search-for” and “difficult-to-search-for” topics.

TARCiS recommendations 4 to 8 comprise guidance for technical aspects of citation searching. This includes selection of seed references, use of electronic citation indexes, deduplication, and iterative citation searching. While composing these recommendations, the TARCiS study group has taken into account that individual workflows must be framed in line with institutional licenses for subscription-only databases and software. For illustration, one such workflow that is based on the licenses as provided by the University of Basel was deposited as an online video.^54^

Concerning guidance for reporting of citation searching, we developed a consensus terminology set for citation searching methods (TARCiS recommendation 1) as well as a recommendation for preferred reporting items for citation searching (TARCiS recommendation 10) along with a downloadable checklist.^17^ The latter exceeds the reporting standards provided by PRISMA-S.^6^ Since the TARCiS and PRISMA-S study groups have some overlap, we suggest that systematic reviewers, methodologists, journal reviewers and editors use TARCiS recommendation 10 as an additional checklist until future work by the PRISMA-S study group produces an updated reporting guideline.

### Dissemination

TARCiS is intended to be used by researchers, systematic reviewers, information specialists, librarians, editors, peer reviewers, and others who are conducting or assessing citation searching methods.

To enhance dissemination among these stakeholders, we aim to provide additional open access publications in scientific and non-scientific journals relevant in the field of information retrieval and evidence syntheses.

We will launch a TARCiS website and plan to announce the TARCiS statement on various platforms (including EQUATOR and PRSIMA). We aim to make it available via the LIbrary of Guidance for HealTh Scientists (LIGHTS), a living database for methods guidance,^55^ The Systematic Review Toolbox, an online catalog of tools for evidence syntheses,^56^ and ResearchGate, a social scientific network to share and discuss publications.

We will announce the TARCiS statement to editors of journals relevant in the field of information retrieval and evidence syntheses to seek their endorsement. This will commit authors and peer-reviewers to use TARCiS to inform their conduct, reporting, and evaluation of citation searching. We will also actively seek endorsement by primary teaching stakeholders in evidence syntheses and systematic literature searching (e.g., York Health Economics Consortium, RefHunter, Cochrane, Joanna Briggs Institute, and the Campbell Collaboration).

We will present and discuss the TARCiS statement on international conferences and share our publications and presentations via relevant mailing lists and newsletters, X (formerly Twitter), and LinkedIn.

### Limitations

A limitation of the TARCiS statement is that, despite the expectation and intent to recruit panelists from all parts of the world, their location was limited to Australia, Europe, and North America. In addition, only a few panelists were recruited from countries in which English was not the dominant language. Furthermore, both the evidence collected in our scoping review and the participating panelists are primarily involved with health-related research. These considerations may reduce the generalizability of our recommendations and research priorities to other countries, languages, and research areas.

## CONCLUSIONS

TARCiS comprises ten specific recommendations on when and how to conduct and report citation searching in the context of systematic literature searches, and four research priorities. It contributes to a unified terminology, systematic application, and transparent reporting of citation searching and will support researchers, systematic reviewers, information specialists, librarians, editors, peer reviewers, and others who are conducting or assessing citation searching methods. In addition, TARCiS may inform future methodological research on the topic. We encourage systematic reviewers and information specialists to incorporate TARCiS into their standardized workflows.

## DECLARATIONS

### Availability of data and materials

The survey sheets and questionnaires that were used for this study are included in the supplementary content. Data generated and analyzed during this study (except sociodemographic information) is available on Open Science Framework (OSF; https://osf.io/y7kh3).

### Ethical approval

Our study is based on published information and uses surveys of topical experts and therefore did not fall under the regulations of the Swiss Human Research Act, and we did not need to apply for ethical approval according to Swiss law. Data protection and privacy issues for the survey are outlined in the main text.

## Acknowledgement

We thank Jill Hayden (Dalhousie University) and Claire Duddy (United Kingdom) for participating in our Delphi panel. We thank Christian Buhtz (Martin Luther University Halle-Wittenberg), Jasmin Eppel-Meichlinger (Karl Landsteiner University of Health Sciences), Tania Rivero (University of Berne), and Monika Wechsler (University of Basel) for participating in the pretest of the Delphi survey.

## Authors’ contributions

All authors made 1) substantial contributions to conception and design, or acquisition of data, or analysis and interpretation of data, 2) drafted the article or revised it critically for important intellectual content, and 3) finally approved the version to be published.

## Core authors’ contributions

The core authors had full access to all of the data in the study and take responsibility for the integrity of the data and the accuracy of the data analysis.

Concept and methodology: Hirt, Nordhausen, Fuerst, Ewald, Appenzeller-Herzog

Acquisition, analysis, interpretation, and validation of data: Hirt, Nordhausen, Fuerst, Ewald, Appenzeller-Herzog

Drafting of the manuscript: Hirt, Appenzeller-Herzog

Critical revision of the manuscript for important intellectual content: Hirt, Nordhausen, Fuerst, Ewald, Appenzeller-Herzog

Statistical analysis: Hirt, Ewald, Appenzeller-Herzog Visualization: Hirt, Nordhausen, Ewald, Appenzeller-Herzog Obtained funding: Not applicable

Administrative, technical, or material support: Hirt, Appenzeller-Herzog Supervision: Hirt, Appenzeller-Herzog

The TARCiS study group authors’ contributions: The study group authors are the Delphi panelists who were involved in Delphi rounds 1 to 4. They received the final manuscript draft for critical revision, important intellectual input, and approval for publication.

---

**The TARCiS statement: Guidance on terminology, application, and reporting of citation searching**

**APPENDIX 1. Consensus ratings of recommendations and research priorities through the Delphi rounds**

## APPENDIX 2. Survey instrument Delphi rounds 1-4

*The TARCiS survey instrument is separated into multiple survey parts (2-6, depending on the Delphi round). Throughout the four Delphi rounds, we labelled the survey parts as Part 1, 2, etc. In the main manuscript, we renamed them to Part A, B, etc. Thus, the labels of the survey parts presented in this appendix deviate from the labels presented in the manuscript (Part 1 corresponds to Part A, etc.). Also, aspects needing further research were renamed to research priorities for the reporting of our findings in the main manuscript*.

**DELPHI SURVEY ROUND 1**

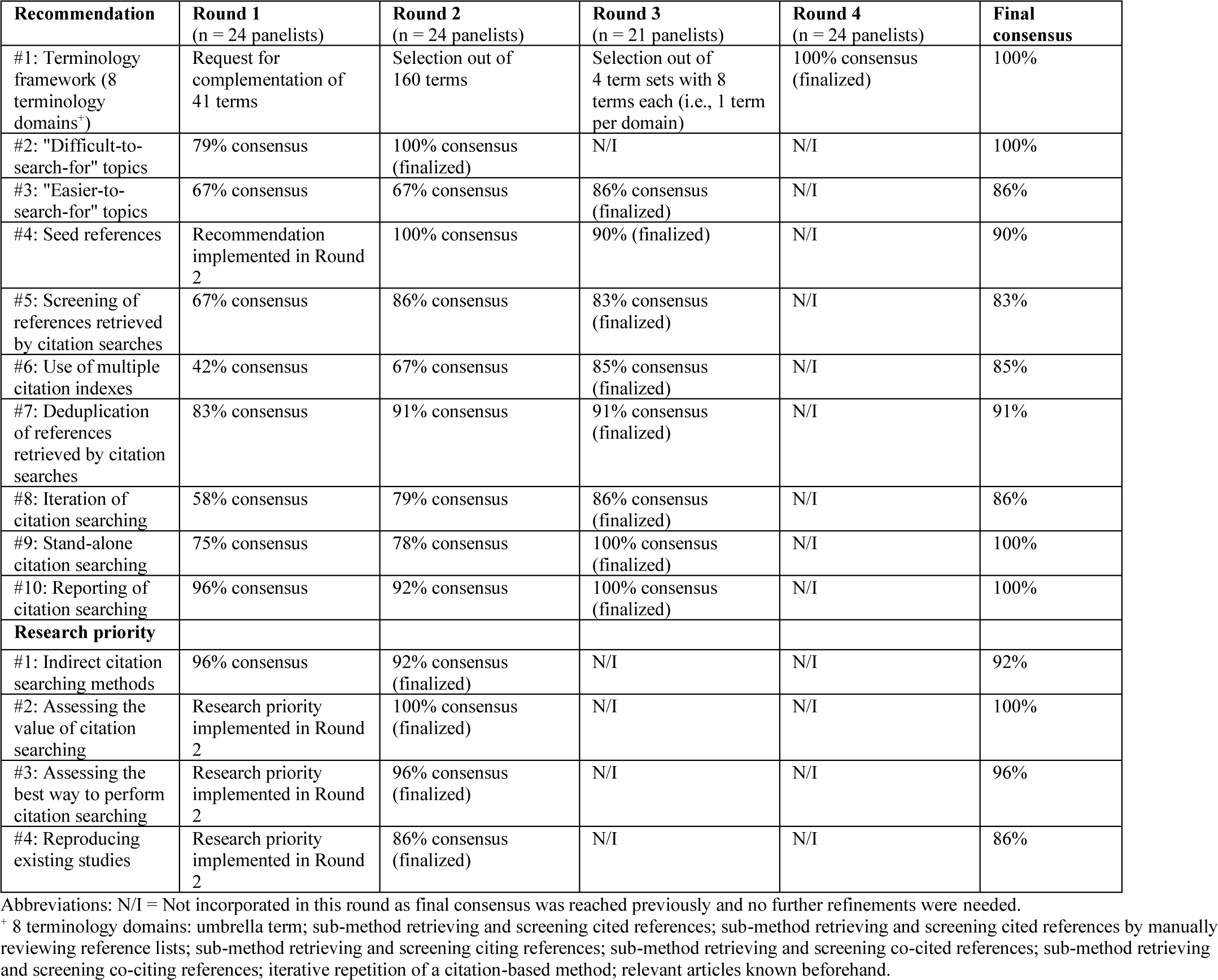

**Part 1: Terminology framework - term collection**

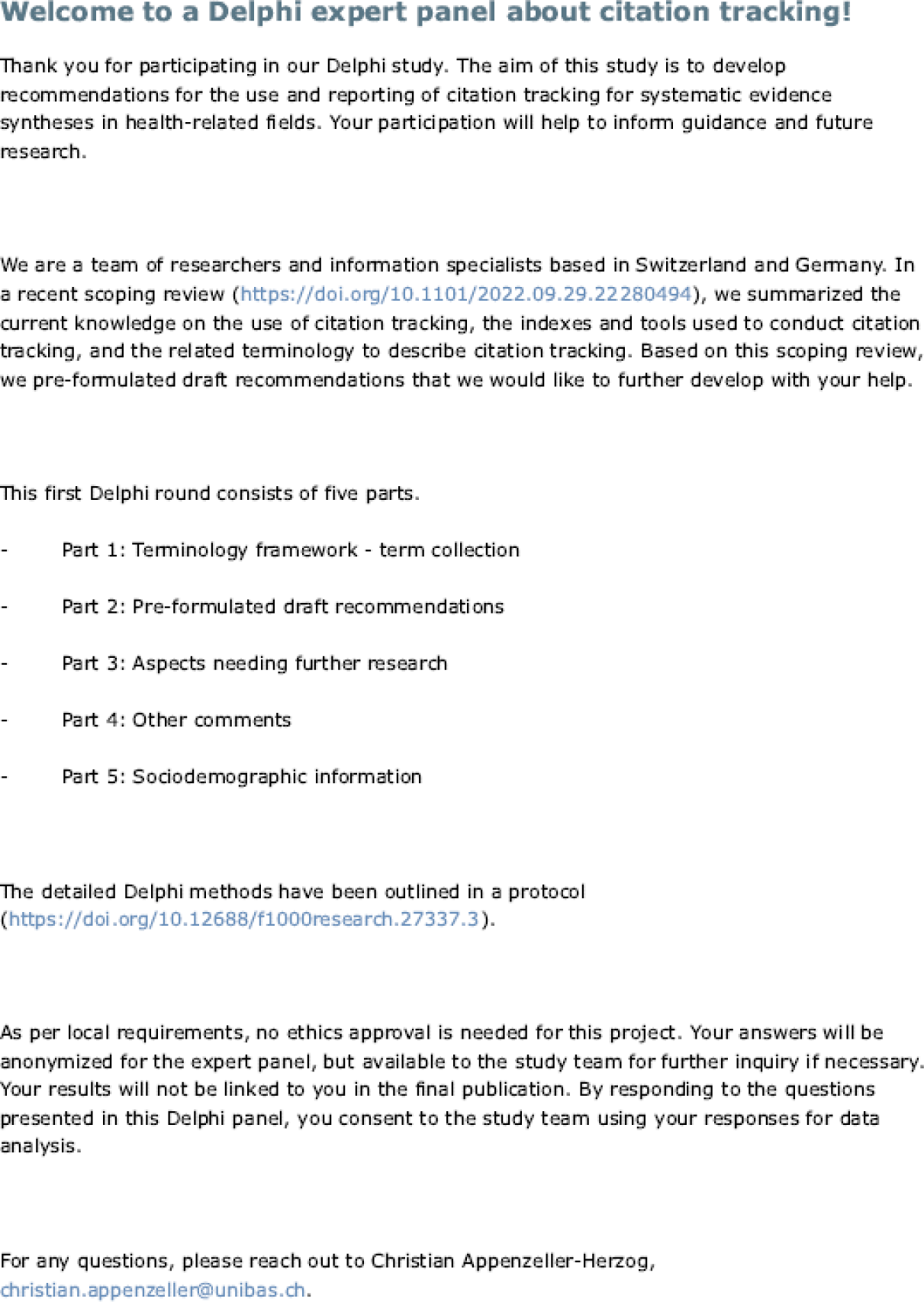

**Part 2: Pre-formulated draft recommendations**

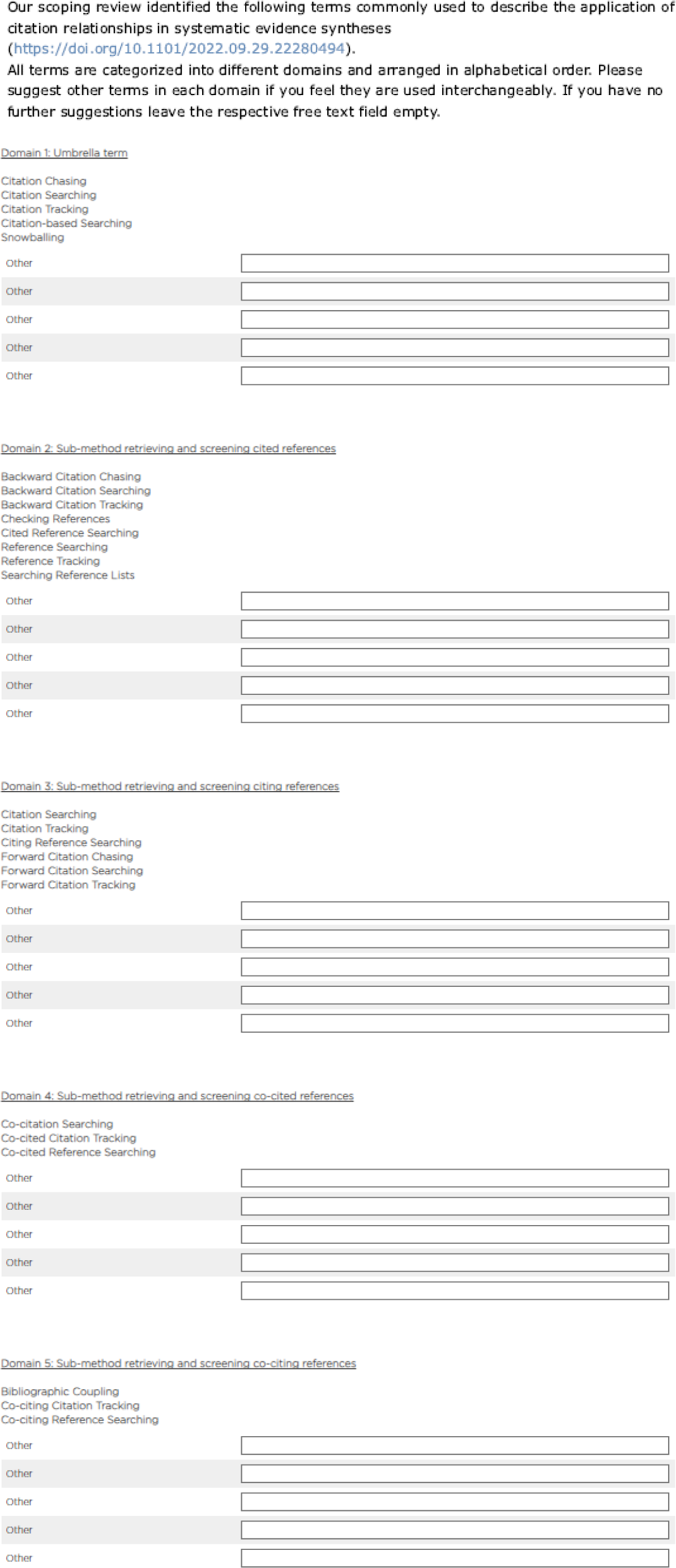
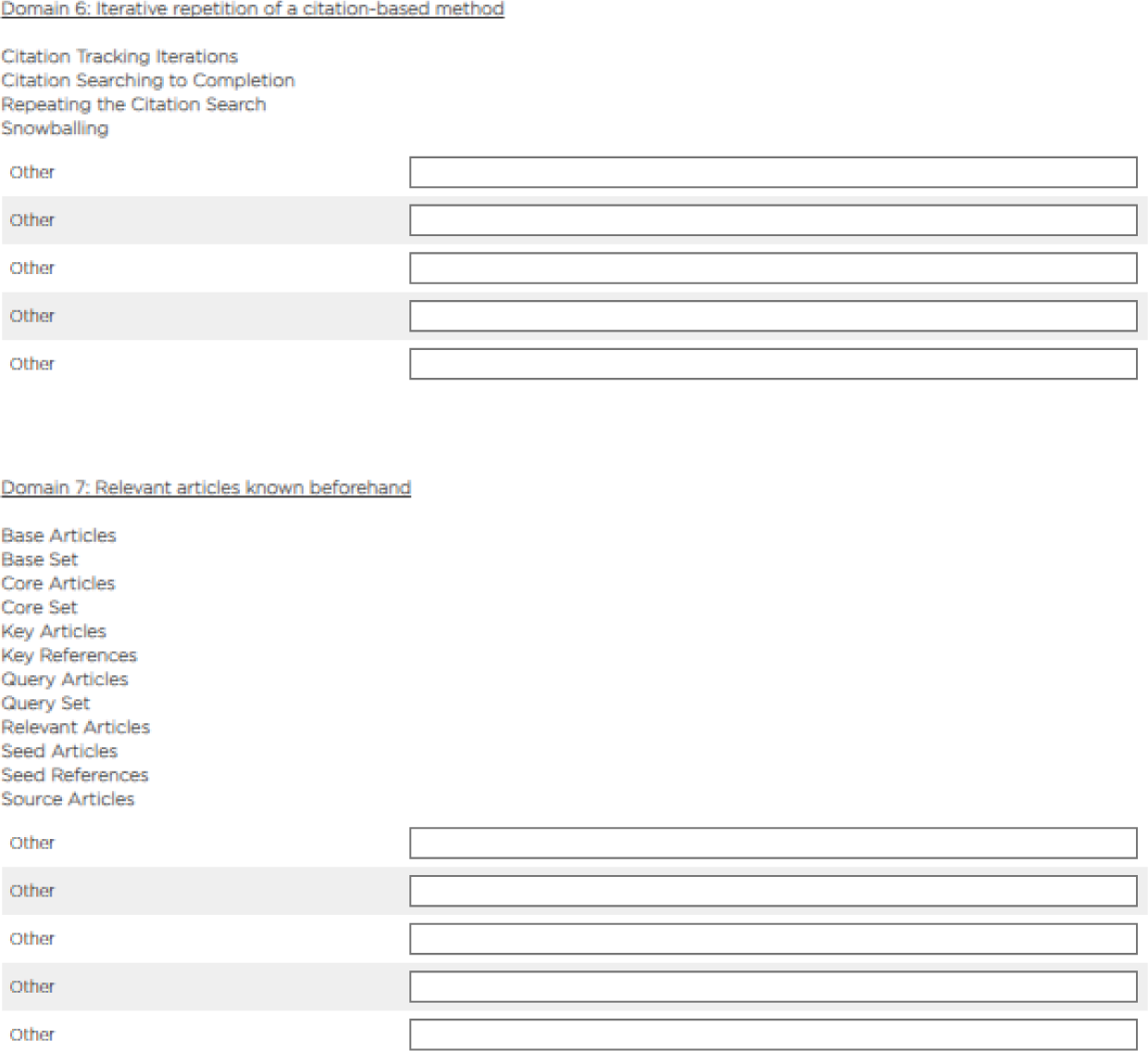

Recommendation 1

**For “hard-to-search-for” systematic search topics, backward and forward citation tracking should be conducted as a supplementary search using all included records from database searching as seed references.**

Rationale: Evidence indicates that the search topic matters as to the question whether citation tracking as a supplementary search technique is likely to add missed references to a systematic literature search. Thus, searches for particular study designs (observational, prognostic, or diagnostic test studies) or health science topics such as non-pharmacological, non-clinical, public health, policymaking, or alternative medicine have been brought into connection with effective supplementary citation tracking searches. The underlying reasons are manifold and include poor amenability of the topic to text-based searching (e.g., owing to inconsistent terminology or vocabulary overlaps with other topics), paucity of research works so that the missing of relevant studies has a higher impact, and lack of prominence of a topic that is predominantly presented in journals that are not indexed in major databases. However, a clean categorization of “hard-to-search-for” topics is currently not possible and it remains upon the review authors themselves to judge whether their review topic is likely to fall into this category.

Recommendation 2

**For “easier-to-search-for” systematic search topics, backward and forward citation tracking using all included records from database searching as seed references may or may not be conducted as a supplementary search.**

Rationale: Evidence indicates that the search topic matters as to the question whether or not citation tracking as a supplementary search technique is likely to add missed references to a systematic literature search. Thus, searches for clearly defined clinical interventions as part of Participant-Intervention-Comparison-Outcome (PICO)-questions have been brought into connection with less effective supplementary citation tracking searches, especially when the search strategies are comprehensive (e.g., covering many databases). However, a clean categorization of “easier-to-search-for” topics is currently not possible and it remains upon the review authors themselves to judge whether their review topic is likely to fall into this category.

Recommendation 3

**Backward citation tracking should be conducted by reviewing the titles and abstracts of the records as provided by a citation index rather than by title alone as provided when reviewing reference lists of the records.**

Rationale: Citation tracking workflows encompass two consecutive steps: retrieval of references and screening of retrieved references for eligibility. When using an electronic citation index for citation tracking, retrieval and screening are usually separated. While forward citation tracking always relies on a citation index, backward citation tracking can also be performed by manually checking the reference lists of the seed papers. Such reference list checking has three disadvantages which may lead to hampered comparability of the results of backward and forward citation tracking: (i) The retrieval and screening of references are no longer separated, (ii) the citation tracking results cannot be deduplicated against each other and/or against the primary search results (see recommendation 5), and (iii) the screening is mainly based on the title field and abstracts cannot be used for eligibility assessments.

Recommendation 4

**Backward and forward citation tracking results should be collected from at least two citation indexes to enhance coverage of eligible references and their citations.**

Rationale: As with electronic databases, citation indexes do not offer 100% coverage. That means that one single citation index does not always cover the whole set of eligible references/seed references and it very likely will not find all the citing and cited literature of these eligible references/seed references. Evidence indicates that when using more than one citation index for citation tracking, the results of the different indexes complement each other. Thus, retrieval of backward and forward citation tracking results from more than one citation index (e.g., TheLens via citationchaser, Scopus, Web of Science) followed by deduplication (see recommendation 5) is a powerful method to increase the sensitivity of citation tracking. It is reminiscent of the complementary effect of using multiple electronic databases for the primary database search, which is the gold-standard in systematic search workflows.

Recommendation 5

**Before screening, collected supplementary backward and forward citation tracking results should be deduplicated against each other and against the primary search results by using a suitable method of deduplication.**

Rationale: The concept of citation tracking as a supplementary search method relies on the notion that reference and cited-by lists of eligible references are topically related to these references. This implies a considerable degree of overlap within these lists. What is more, the overlap likely also extends to the results of the primary systematic search that was performed on the same topic. Based on these considerations and on the fact that the results of the primary search have already been screened for eligibility, the screening load of citation tracking results can be significantly cut by removing those references that have already been screened for eligibility and those that appear as duplicates in more than one reference or cited-by lists.

Recommendation 6

**If the screening of citation tracking results leads to the identification of additional eligible records, another iteration of citation tracking using these records as seed references should be conducted.**

Rationale: Using newly retrieved, relevant references as new seed references is referred to as citation tracking iterations. Citation tracking methods can be conducted over one or more iteration(s). There is evidence that conducting citation tracking iterations can contribute to the comprehensiveness of a systematic search. Very much like what is written in the explanatory text to recommendation 5, the results of citation tracking iterations should be thoroughly deduplicated to reduce the screening load.

Recommendation 7

**For literature search projects that do not aim at completeness of recall, stand-alone direct and/or indirect citation tracking methods can be considered, but stand-alone citation tracking should not be used for searches that aim at completeness of recall.**

Rationale: Traditionally, backward citation tracking and, more lately, also forward citation tracking are being used as supplementary search techniques to retrieve and add missed references to a primary systematic database search. We refer to stand-alone citation tracking when citation tracking is used as the primary search method without prior database searching. Seed references for stand-alone citation tracking could be references from private collections or retrieved from simplified database searches. To enhance recall, stand-alone citation tracking can also, in addition to backward and forward citation tracking, involve indirect methods that collect co-citing and co-cited references. When study authors replicated published systematic reviews with stand-alone citation tracking, they rarely retrieved 100% of the literature that was included in the systematic review. Since search methods for systematic reviews and scoping reviews should aim at completeness of recall, stand-alone citation tracking is not a suitable method for these types of literature review.

Recommendation 8

**In addition to the guidance PRISMA-S already provides, reporting of citation tracking should clearly state**

- **the date(s) of searching (which may differ between citation tracking iterations),**
- **the number of citation tracking iterations,**
- **all citation indexes searched (e.g., Scopus, Web of Science, Google Scholar) and, if applicable, the tools that were used to access them (e.g., Publish or Perish, citationchaser),**
- **the method of deduplication, and**
- **the method of screening (e.g. statement if the same method as for the primary search results was applied or explanation of what was done if it deviated).**

Rationale: The relevant guidance for researchers conducting citation tracking in systematic literature searching can be found in item 5 “citation searching” of PRISMA-S (https://doi.org/10.5195/jmla.2021.962). According to PRISMA-S, required reporting items are the directionality of citation tracking (examination of cited or citing references), methods and resources used for citation tracking (bibliographies in full text articles or citation indexes), and the seed references that citation tracking was performed upon. Other important elements of citation tracking that emerged from our scoping review (https://doi.org/10.1101/2022.09.29.22280494), however, remained neglected or should be specified. These elements are listed in recommendation 8 as a supplement to PRISMA-S to comprehensively guide the reporting of supplementary citation tracking in systematic literature searching.

Other recommendations

**Part 3: Aspects needing further research**

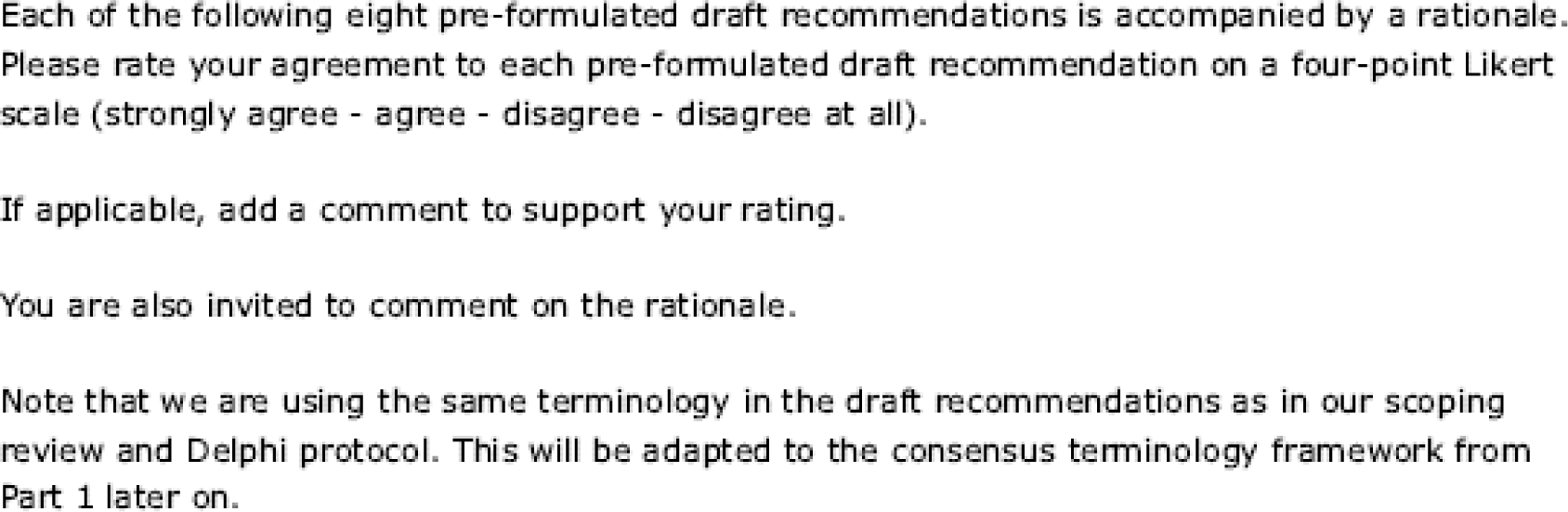

Aspect 1

**Development of recommendations for applicability and conduct of indirect citation tracking methods as supplementary methods in systematic searching (including possible prioritization and cut-off of results) requires further research.**

Rationale: Indirect citation tracking involves the collection and screening for eligibility of references that share references in their bibliography or citations with one of the seed references (i.e., co-citing or co-cited references). Indirect citation tracking typically retrieves a large volume of records to be screened. Therefore, prioritization algorithms and cut-offs have been proposed that aim at reducing the workload of eligibility screening. The methodological studies that so far have pioneered indirect citation tracking methods for health-related topics have exclusively focused on stand-alone citation tracking. Therefore, the judgement of the potential utility of indirect citation tracking methods as supplementary search techniques in systematic reviews requires further methodological research.

Other aspects

**Part 4: Other comments**

**Do you have any other gerneral comments to share? If so, please provide them here.**

**Part 5: Sociodemographic information**

**We ask you for sociademographic information to summarize key characteristics for the panel.**

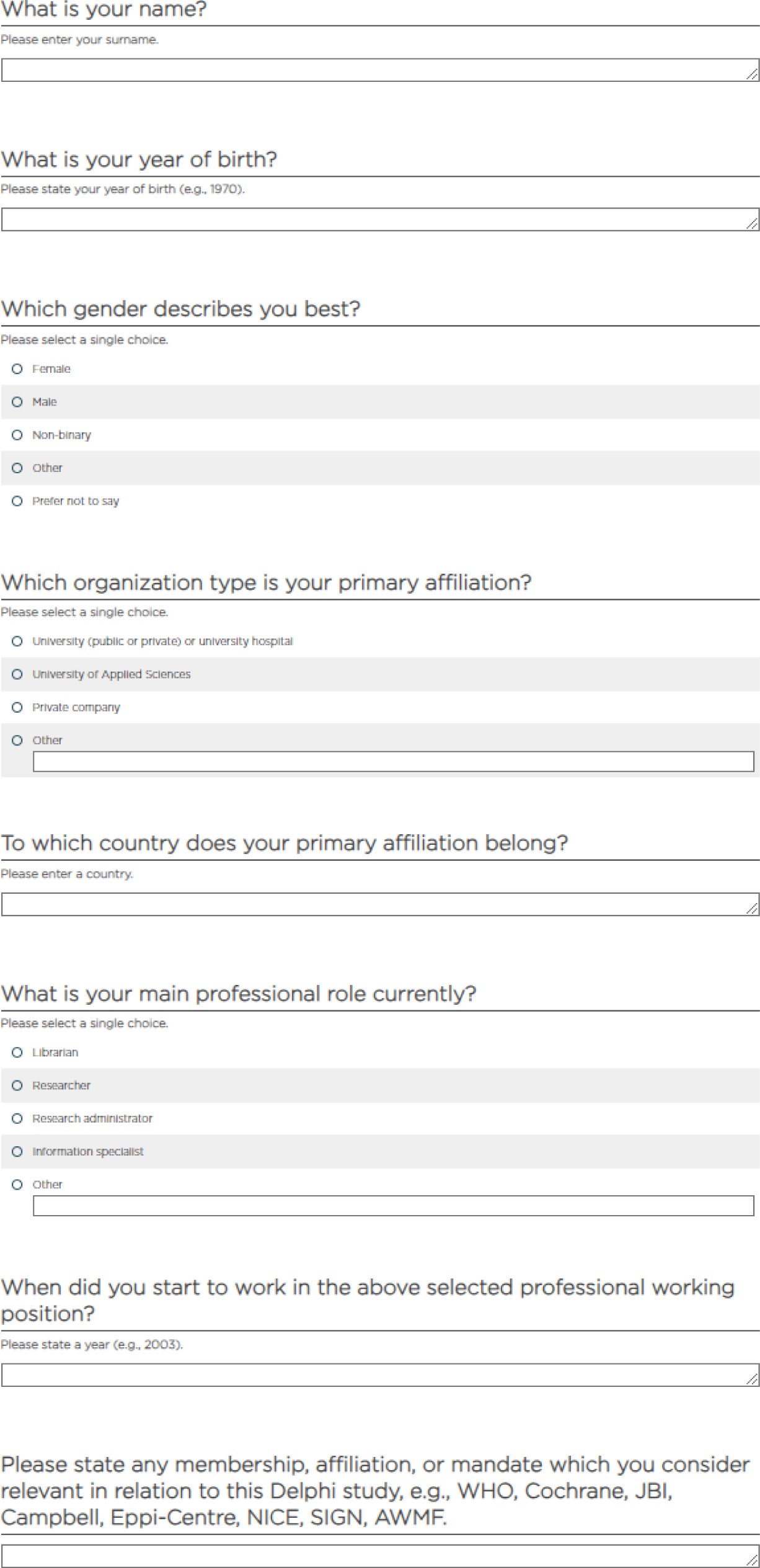

**DELPHI SURVEY ROUND 2**

Welcome to round 2 of our Delphi study and thank you for participating!

The aim of this study is to develop recommendations for the use and reporting of citation tracking for systematic evidence syntheses in health-related fields. Your participation will help to inform guidance and future research.

This second Delphi round consists of six parts.

Part 1: Terminology framework - term selection

Part 2: Amended draft recommendations

Part 3: New draft recommendations based on your input

Part 4: Amended aspects needing further research

Part 5: New aspects of citation tracking needing further research based on your input

Part 6: Other comments

For your information, we prepared a <u>document detailing the changes compared to the first Delphi round</u>. In this document, we also outline why these changes were done and the percentage of agreement for the original recommendation as achieved in round 1. Please also find a figure <u>here</u> that describes the terminology used in this Delphi. The Delphi methods have been outlined in a <u>peer-</u> <u>reviewed protocol</u>, and details have been pre-specified in an internal protocol published on <u>Open Science Framework</u>.

As per local requirements, no ethics approval is needed for this project. Your answers will be anonymized for the expert panel, but available to the study team for further inquiry if necessary. They will not be linked to you in the final publication. By answering to the questions presented in this Delphi round, you consent to our use of your answers for data analysis.

For any questions, please reach out to Christian Appenzeller-Herzog.

**Part 1: Terminology framework - term selection**

Please find the list with the suggested terms for the different terminology domains arranged in alphabetical order.

**Terminology Domain 1: Umbrella Term**

Please select your preferred 1 to 3 items.

Citation analysis

Citation-based searching

Citation chaining Citation chasing Citation following

Citation network searching Citation path

Citation searching Citation spidering Citation tracing Citation tracking Cited-by search Cited citations Cited Records Pearl-growing Pearling

Record analysis

Record-based searching Record chaining Record chasing

Record following

Record network searching Record path

Record searching Record spidering Record tracing Record tracking Reference chasing Reference checking Reference tracking

Snowballing

Using citation alerts

**Terminology Domain 2: Sub-method retrieving and screening cited references**

Please select your preferred 1 to 3 items. Backward citation chaining

Backward citation chasing

Backward citation following

Backward citation searching

Backward citation tracking

Bibliography trawling

Checking bibliographies

Checking reference lists

Checking references

Cited reference check

Cited reference searching

Cited references

Citing work

Cross-citations

Footnote chasing

Reference checking

Reference harvesting

Reference list screening

Reference searching

Reference tracking

Review reference lists

Reviewing bibliographies

Screening bibliography

Screening references

Searching reference lists

**Terminology Domain 3: Sub-method retrieving and screening citing references**

Please select your preferred 1 to 3 items.

Citation checking

Citation searching

Citation tracking

Cited by searching

Citing reference check

Citing reference searching

Finding citing articles

Forward citation chaining

Forward citation chasing

Forward citation following

Forward citation searching

Forward citation tracking

**Terminology Domain 4: Sub-method retrieving and screening co-cited references**

Please select your preferred 1 to 3 items.

Backward co-citation tracking

Co-citation analysis

Cocitation database search

Co-citation searching

Co-citation tracking

Co-cited citation tracking

Co-cited reference searching

CoCites searching

Companion-citation searching

Companion-citation tracking Companion-paper searching Companion-paper tracking

**Terminology Domain 5: Sub-method retrieving and screening co-citing references**

Please select your preferred 1 to 3 items.

Bibliographic coupling

Co-citing analysis

Co-citing citation tracking

Co-citing reference searching

Companion reference

Forward co-citation tracking

Related records

**Terminology Domain 6: Iterative repetition of a citation-based method**

Please select your preferred 1 to 3 items.

Citation chaining

Citation mining

Citation searching to completion

Citation tracking iterations

(Co-)Citation searching to saturation Cycling

Iterative citation analysis

Iterative citation searching

Multiple rounds of citation searching

Multi-step citation searching n-degree citation search (where n = number of iterations)

Pearl growing

Pearling

Phased citation searching

Repeated/Exhausted until there are no new items

Repeating the citation search

Snowballing

**Terminology Domain 7: Relevant articles known beforehand**

Please select your preferred 1 to 3 items.

Base articles

Base documents

Base papers

Base references

Base set

Benchmarking articles

Benchmarking documents

Benchmarking set

Benchmarking studies

Central articles

Core articles

Core documents

Core papers

Core references

Core set

Crucial articles

Design set of articles

Desk draw set

Development set of articles

Eligible articles

Eligible studies

Filedraw set

Included articles

Included studies

Key articles

Key documents

Key papers

Key references

Known articles

Known documents

Known includes known relevant references

Known studies

Leading articles

Major references

Original articles

Query articles

Query set

Reference set

Relevant articles

Relevant documents

Relevant papers

Research team’s library

Research team’s reference library

Researchers’ library

Researchers’ reference library

Seed articles

Seed papers

Seed references Seeds

Seminal articles

Source articles

Source documents

Source papers

Starting articles Test set

**Part 2: Amended draft recommendations**

We amended the recommendations according to the feedback of the Delphi panel. Concerning some recommendations, there appear to be “fractions” with different opinions in the panel, e.g., regarding the use of automated or manual backward citation tracking. Please note that we have tried to amend the recommendations to achieve the broadest possible consensus.

Please also note that as per your input, we have phrased a new Recommendation 3 on the selection of seed references. At the same time, the former statements regarding seed references have been removed from Recommendations 1 and 2.

The rationale texts are currently lacking referencing as all the underlying evidence is cited and synthesized in the Scoping review we have shared with you before the Delphi procedure (https://doi.org/10.1101/2022.09.29.22280494). We are going to reference everything in detail in the final publication, except for aspects that are based on expert opinions from this Delphi for which no evidence is available.

Recommendation 1

**For “hard-to-search-for” systematic search topics, backward and forward citation tracking should be seriously considered as supplementary search techniques.**

Rationale: Evidence indicates that the ability of citation tracking as a supplementary search technique to find additional unique records in a systematic literature search varies with the topic. Searches for particular study designs (qualitative, mixed-method, observational, prognostic, or diagnostic test studies) or health science topics such as non-pharmacological, non-clinical, public health, policymaking, service delivery, or alternative medicine have been linked with effective supplementary citation tracking searches. The underlying reasons are manifold and include poor transferability of the topic to text-based searching (e.g., owing to poor conceptual clarity, inconsistent terminology, or vocabulary overlaps with other topics). The ability of citation tracking to find any publication type including unpublished or grey literature or literature that is not indexed in major databases (e.g., concerning a developing country) may also be relevant. However, a clean categorization of “hard-to-search-for” topics is currently not possible and it remains for the review authors themselves to judge whether their review topic is likely to fall into this category.

Independent of how hard or difficult a topic is to search for, researchers with limited experience in systematic searching, with uncertainty that their strategy comprehensively captures the topic, and without help from an experienced information specialist may also take citation tracking into consideration.

Recommendation 2

**For “easier-to-search-for” systematic search topics and when a text-based search is highly sensitive, formal backward and forward citation tracking are not explicitly recommended as supplementary search techniques. Screening the reference lists of included records (manual backward citation tracking) is always an option.**

Rationale: Evidence indicates that the ability of citation tracking as a supplementary search technique to find additional unique references in a systematic literature search varies with the search topic. Searches for clearly defined clinical interventions as part of Participant-Intervention-Comparison-Outcome (PICO)-questions have been linked with less effective supplementary citation tracking searches, especially when the search strategies are sensitive and conducted in several databases. However, a clean categorization of “easier-to-search-for” topics is currently not possible and it remains for the review authors themselves to judge whether their review topic is likely to fall into this category. As manual backward citation tracking is still part of the workflow of many researchers, we do not intend to discourage this procedure. However, depending on the overall number of included records and backward references, this may be more time-consuming (number of duplicates) and less thorough (only titles and context available) than automated approaches with citation indexes and/or citation tracking tools. Independent of how hard or difficult a topic is to search for, researchers with limited experience in systematic searching, with uncertainty that their strategy comprehensively captures the topic, and without help from an experienced information specialist may also take citation tracking into consideration.

Recommendation 3

**Backward and forward citation tracking as supplementary search techniques should be based on all included records of the primary search, i.e., all records that meet the inclusion criteria of the review after screening. Depending on the situation, using further pertinent records as additional seed references, or using only a defined sample of the included records may be considered. If not all included records are used for citation tracking, the selection of seed references should be justified.**

Rationale: The more seed references are used, the better are the chances that citation tracking finds additional relevant unique records. While using only a sample of the included records as seed references may be enough, there is currently no evidence that could help decide how many seeds are needed or how to decide which may perform better. Hence, we recommend using all the records that meet the inclusion criteria of the review. However, if a review deals with a very small or large number of included records, using all of these as seed references may be too little helpful or too work intense, respectively. In such situations, it can be taken into consideration to also use other pertinent records such as systematic reviews on the topic that were flagged during the screening phase or to use a restricted sample of included records, e.g., the 30 most cited records.

Recommendation 4

**Backward citation tracking should ideally be conducted by screening the titles and abstracts of the records as provided by a citation index. Screening titles as provided when manually reviewing reference lists of the included records can be performed as an add-on.**

Rationale: Citation tracking workflows encompass two consecutive steps: retrieval of records and screening of retrieved records for eligibility. When using an electronic citation index for citation tracking, retrieval and screening are usually separated. While forward citation tracking requires a citation index, backward citation tracking can also be performed by manually checking the reference lists of the seed references. Manual backward citation tracking is sometimes part of an established workflow, e.g., done during eligibility-checking of the full-text record or during data extraction. Merging these two steps has the benefit that researchers know the context in which a reference was used and that all references can be screened. However, manual backward citation tracking has three disadvantages: (i) The retrieval and screening phases are no longer separated which makes reporting of the methods/results difficult and unclear, (ii) the citation tracking results cannot be deduplicated against each other and/or against the primary search results which may add an unnecessarily high workload (see recommendation 6), and (iii) the eligibility assessments are restricted to the titles (instead of titles and abstracts) which could lead to relevant records being overlooked due to unspecific titles mentioned in vague contexts.

In recent years, online citation tracking options via citation indexes or free to access citation tracking tools have become more readily available with faster and easier procedures. More and even better tools to facilitate this workflow are expected in the future. Combining citation tracking via citation indexes with automated deduplication (free online tools available) makes this recommendation feasible. On the other hand, citation indexes will in most cases fail to retrieve all the cited references. Thus, references to grey literature (such as websites or registry entries) or poorly indexed literature may only be retrievable by manual backward citation tracking or by a subset of citation indexes.

Recommendation 5

**Collection of backward and forward citation tracking results from at least two citation indexes should be considered if access is available and, especially, when the coverage of seed references and their citations is low.**

Rationale: A single citation index may not cover all seed references and it very likely will not find all the citing and cited literature. The reason for that is that citation indexes do not offer 100% coverage as some references are currently not indexed in one or several citation index(es). Evidence indicates that when using more than one citation index for citation tracking, the results of the different indexes complement each other. Thus, retrieval of backward and forward citation tracking results from more than one citation index (e.g., TheLens via citationchaser, Scopus, citation indexes in Web of Science) followed by deduplication (see recommendation 6) is a powerful method to increase the sensitivity of citation tracking. It is reminiscent of the complementary effect of using multiple electronic databases for the primary database search, which is the gold-standard in systematic search workflows. In recent years, online citation tracking options have increased many of which providing open access tools and indexes that make electronic citation tracking universally accessible, faster, and easier. More and even better tools to facilitate citation-index-based workflows are expected in the future.

Recommendation 6

**Before screening, the results of supplementary backward and forward citation tracking should be deduplicated against each other and the primary search results.**

Rationale: The concept of citation tracking as a supplementary search method relies on the notion that reference and cited-by lists of eligible references are topically related to these references. This implies a considerable degree of overlap within these lists. What is more, the overlap likely also extends to the results of the primary systematic search that was performed on the same topic. Based on these considerations and on the fact that the results of the primary search have already been screened for eligibility, the screening load of citation tracking results can be significantly cut by removing those references that have already been screened for eligibility and those that appear as duplicates in more than one reference or cited-by lists.

There are several methods of deduplication. While it can be conducted manually, nowadays standard bibliographic management software and specialized tools provide automated deduplication solutions, allowing for easier and faster processing.

If citation tracking leads to only a small number of results, omission of the deduplication step can be considered to save time and administrative effort.

Recommendation 7

**If citation tracking finds additional eligible records, another iteration of citation tracking should be considered using these records as new seed references.**

Rationale: Using newly retrieved, relevant references as new seed references is referred to as citation tracking iterations. Citation tracking methods can be conducted over one or more iteration(s). There is evidence that conducting citation tracking iterations can contribute to the comprehensiveness of a systematic search.

Since further iterations require additional time and effort and may interrupt the ongoing review process, the decision in favor of or against further iterations should be guided by a cost-benefit assessment. Relevant factors to be assessed include the review topic (hard-or easy-to-search-for), sensitivity of the primary search, aim for completeness of the literature search, and the estimated potential benefit of the iteration(s) (e.g., based on the number/percentage of included records found with the previous citation tracking).

As outlined in the rationale of recommendation 6, results of citation tracking iterations can be deduplicated against all previously retrieved records to reduce the screening load.

Recommendation 8

**Stand-alone citation tracking should not be used for literature searches that aim at completeness of recall. If the search does not aim at completeness, stand-alone citation tracking can be considered.**

Rationale: We refer to stand-alone citation tracking when any form of citation tracking is used as the primary search method without prior database searching, contrary to its traditional use as a supplementary search method to a primary database search. Seed references for stand-alone citation tracking could be records from private collections or retrieved from non-systematic literature searches. To enhance recall, stand-alone citation tracking can also, in addition to backward and forward citation tracking, involve indirect methods that collect co-citing and co-cited references. When study authors replicated published systematic reviews with stand-alone citation tracking, they mostly missed literature that was included in the systematic review. Since search methods for systematic reviews and scoping reviews should aim at completeness of recall, stand-alone citation tracking is not a suitable method for these types of literature review.

Recommendation 9

**In addition to the guidance PRISMA-S already provides, reporting of citation tracking should clearly state**

-**the date(s) of searching (which may differ between citation tracking iterations) (not applicable for manual backward citation tracking),**

-**the number of citation tracking iterations,**

-**all citation indexes searched (e.g.,** LENS.ORG**, Google Scholar, Scopus, citation indexes in Web of Science) and, if applicable, the tools that were used to access them (e.g., Publish or Perish, citationchaser),**

-**information about the duplication process (e.g., manual/automated, the software or tool used), and**

-**the method of screening (e.g., statement whether the records were screened in the same way as the primary search results or description of the alternative method used).**

Rationale: The relevant guidance for researchers conducting citation tracking in systematic literature searching can be found in item 5 “citation searching” of PRISMA-S (https://doi.org/10.5195/jmla.2021.962). According to PRISMA-S, required reporting items are the directionality of citation tracking (examination of cited or citing references), methods and resources used for citation tracking (bibliographies in full text articles or citation indexes), and the seed references that citation tracking was performed upon. While PRISMA-S can be seen as minimum reporting standard for citation tracking as supplementary search technique, other important elements that emerged from our scoping review (https://doi.org/10.1101/2022.09.29.22280494) need to be reported to achieve full transparency and/or reproducibility. These elements are listed in recommendation 9 as a supplement to PRISMA-S to comprehensively guide the reporting of supplementary citation tracking in systematic literature searching.

Please note that this information does not have to be reported in the main methods of a study. Ideally, there is a comprehensive appendix or link to a repository with all pertinent information regarding the systematic search.

**Part 3: New draft recommendations based on your input**

We have tried to phrase your suggestions as new draft recommendations. Please feel free to make further wording and/or rationale suggestions, especially if you agree with the general recommendation. Should these new recommendations get 75% agreement or more, we will, where sensible, merge them with existing recommendations (they seem to mostly concern the reporting recommendation 9).

For one planned draft recommendation (recommendation 14), you are asked to vote for one of four options before we will phrase a recommendation and rationale text for Delphi Round 3.

Recommendation 10

**Review authors should state who conducted the citation tracking.**

Recommendation 11

**If citation tracking was not conducted, review authors should give a rationale why it was omitted.**

Recommendation 12

**When citation tracking is reported, review authors should state how many and which references were used for the citation tracking.**

*Note: This is already mentioned in the PRISMA-S explanation section. As a recommendation, we would like to improve the visibility*.

Recommendation 13

**The bibliographic details of all CT results should be shared in a file (e.g., a Research Information System (RIS) file via a repository).**

Recommendation 14 (in preparation)

Recommendation on where to report the results of citation tracking in the PRISMA 2020 flow diagram. Please vote for one of the following options:

-It is at the discretion of the authors if they report the results of citation tracking in the left column “Identification of studies via databases and registers” or in the right column “Identification of studies via other methods”. *(current status quo:* https://doi.org/10.5195/jmla.2022.1449*: “users are free to modify the PRISMA 2020 flow diagram templates in a way they consider most optimal for their review”)*

-Results of citation tracking that was done in databases (or in databases via a tool) should be reported in the left column under “Identification of studies via databases and registers” and results of citation tracking that was done manually in the right column “Identification of studies via other methods”, respectively.

-Citation tracking should always be reported in the left column “Identification of studies via databases and registers”, following the rationale that databases will be searched even if for a different purpose.

-Citation tracking as supplementary search method should always be reported in the right column “Identification of studies via other methods”, following the rationale that supplementary search methods should be uniformly reported as “other methods”. Citation tracking as primary search method (i.e., instead of database searching) should be reported in the left column “Identification of studies via databases and registers”.

**Part 4: Amended aspect needing further research**

Please rate your agreement on the amended aspect of citation tracking needing further research.

Aspect 1

**The effectiveness, applicability and conduct of indirect citation tracking methods as supplementary search methods in systematic reviewing require further research (including retrieval of additional unique references, their relevance for the review, and prioritization and cut-off of results).**

Rationale: Indirect citation tracking involves the collection and screening for eligibility of records that share references in their bibliography or citations with one of the seed references (i.e., co-citing or co-cited references). Indirect citation tracking typically retrieves a large volume of records to be screened. Therefore, prioritization algorithms and cut-offs have been proposed that aim at reducing the workload of eligibility screening. The methodological studies that have pioneered indirect citation tracking methods for health-related topics have so far exclusively focused on stand-alone citation tracking. It is currently unclear if the added workload and resources for searching and screening warrant indirect citation tracking methods as supplementary search techniques in systematic reviews of any type (qualitative/quantitative, hard/easy-to-search-for).

**Part 5: New aspects of citation tracking needing further research based on your input**

We are listing a summary of all additional aspects needing further research that have been submitted by the panel, asking for your agreement.

Please only express your agreement, if the aspect (i) is specifically concerned with citation tracking and (ii) you agree it requires future research effort.

Aspect 2

**Further research is needed to assess the value of citation tracking. Potential research topics could be:**

-**influence of citation tracking on results and conclusions of systematic evidence syntheses,**

-**topics or at least determinants of topics where citation tracking likely/not likely has additional value, or**

-**economic evaluation of citation tracking.**

Aspect 3

**Further research is needed to assess the best way to perform citation tracking. Potential research topics could be:**

-**optimal selection of seed references,**

-**optimal use of indexes and tools and their combination to conduct citation tracking,**

-**methods and tools for deduplication of citation tracking results,**

-**subjective influences on citation tracking (e.g., experience of researcher, prevention of mistakes), or**

-**reproducibility of citation tracking.**

Aspect 4

**Further research is needed to reproduce existing studies: Any recommendations in this Delphi that are based on only 1-2 studies require reproduction of these studies in form of larger, prospectively planned studies that grade the evidence for each recommendation and propose additional research where the grade of evidence is weak.**

**Part 6: Other comments**

Provide any other comment as deemed relevant.

**DELPHI SURVEY ROUND 3**

Welcome to round 3 of our Delphi study about citation tracking and thank you for participating!

The aim of this study is to develop recommendations for the use and reporting of citation tracking for systematic evidence syntheses in health-related fields.

**Please note that the focus of this study is not limited to citation tracking used for the review type systematic reviews but concerns any systematic literature searching approach and review type of the systematic evidence synthesis family including those that do not aim at completeness.**

This second Delphi round consists of four parts:

Part 1: Terminology framework - selection of a term set Part 2: Amended draft recommendations

Part 3: Amended aspects needing further research Part 4: Other comments

For your information, we prepared a <u>document detailing the changes compared to the second Delphi round</u>. In this document, we also outline why these changes were done and the percentage of agreement for the original recommendation as achieved in round 2.

Please also find a figure <u>here</u> that describes direct and indirect citation tracking methods. The Delphi methods have been outlined in a <u>peer-reviewed protocol</u>, and details have been pre-specified in an internal protocol published on <u>Open Science Framework</u>.

As per local requirements, no ethics approval is needed for this project. Your answers will be anonymized for the expert panel, but available to the study team for further inquiry if necessary. They will not be linked to you in the final publication. By answering to the questions presented in this Delphi round, you consent to our use of your answers for data analysis.

Finally, we are happy to let you know that the scoping review that has collected the evidence base for this Delphi study has in the meantime been published (https://doi.org/10.1002/jrsm.1635).

For any questions, please reach out to Christian Appenzeller-Herzog.

**Part 1: Terminology framework - selection of a term set**

As per protocol, four harmonized term sets were developed based on the voting results in Delphi round 2.

Outlook: In the final Delphi Round 4, the terminology of the best-ranking term set will be implemented in all recommendation and rationale texts.

Please select your favorite term set.

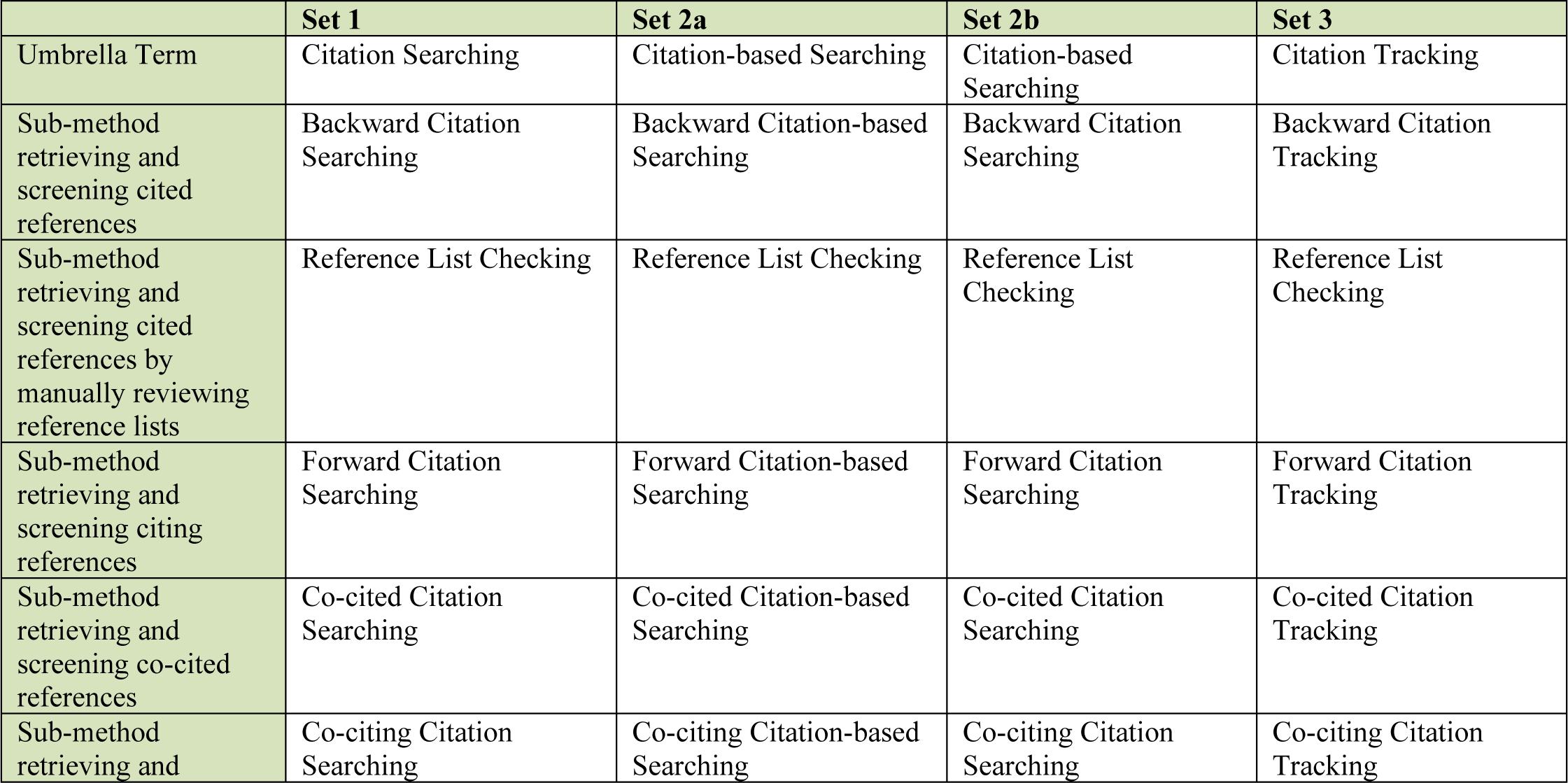
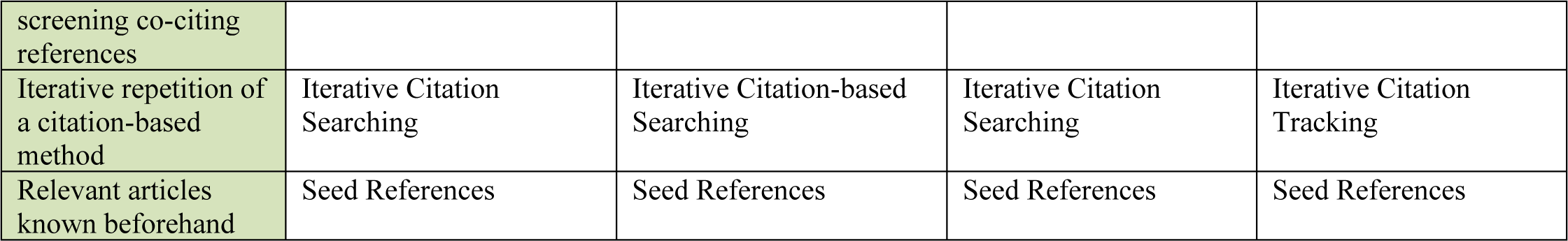

**Part 2: Amended draft recommendations**

We amended the recommendations according to the feedback of the Delphi panel. Recommendations 1, 4 and 6 are now consented (>75% agreement with no relevant changes necessary) and are not to be rated in Delphi round 3 any longer.

The “rationale” headings were rephrased to “rationale and explanatory details”. These texts are currently lacking referencing as all the underlying evidence is cited and synthesized in the Scoping review we have shared with you before the Delphi procedure (https://doi.org/10.1002/jrsm.1635). We are going to reference everything in detail in the final publication, except for aspects that are based on expert opinions from this Delphi for which no evidence is available.

Recommendation 1 (Not included in Delphi round 3)

**For “difficult-to-search-for” systematic search topics, backward and forward citation tracking should be seriously considered as supplementary search techniques.**

Rationale and explanatory details:

Evidence indicates that the ability of citation tracking as a supplementary search technique to find additional unique records in a systematic literature search varies with the topic. Searches for particular study designs (qualitative, mixed-method, observational, prognostic, or diagnostic test studies) or health science topics such as non-pharmacological, non-clinical, public health, policymaking, service delivery, or alternative medicine have been linked with effective supplementary citation tracking searches. The underlying reasons are manifold and include poor transferability of the topic to text-based searching (e.g., owing to poor conceptual clarity, inconsistent terminology, or vocabulary overlaps with other topics). The ability of citation tracking to find any publication type including unpublished or grey literature or literature that is not indexed in major databases (e.g., concerning a developing country) may also be relevant. However, a clear categorization of “difficult-to-search-for” topics is currently not possible and it remains for the review authors themselves to judge whether their review topic is likely to fall into this category.

We recommend that persons conducting the search who have difficulty assessing whether the topic is difficult-or easier-to-search-for always opt for citation tracking or consult an experienced information specialist. If for whatever reason the search strategy does not comprehensively capture the topic, backward and forward citation tracking may compensate the potential loss of information at least to some extent.

Recommendation 2

**For “easier-to-search-for” systematic search topics covered by a highly sensitive search, backward and forward citation tracking are not explicitly recommended as supplementary search techniques. Reference list checking of included records can be used to confirm the sensitivity of the search strategy.**

Rationale and explanatory details:

Evidence indicates that the ability of citation tracking as a supplementary search technique to find additional unique references in a systematic literature search varies with the search topic. Searches for clearly defined clinical interventions as part of Participant-Intervention-Comparison-Outcome (PICO)-questions have been linked with less effective supplementary citation tracking searches, especially when the search strategies are sensitive and conducted in several databases. However, a clear categorization of “easier-to-search-for” topics is currently not possible and it remains for the review authors themselves to judge whether their review topic is likely to fall into this category.

By checking reference lists within the full-texts of seed references, review authors can test the sensitivity of their primary search strategy (i.e. electronic database search). Should no additional relevant, unique studies be found, the primary search may have been sensitive enough. Should additional relevant, unique studies be found, it may be an indication that the primary search was not sensitive enough.

We recommend that persons conducting the search who have difficulty assessing whether the topic is difficult-or easier-to-search-for always opt for citation tracking or consult an experienced information specialist. If for whatever reason the search strategy does not comprehensively capture the topic, backward and forward citation tracking may compensate the potential loss of information at least to some extent.

Recommendation 3

**Backward and forward citation tracking as supplementary search techniques should be based on all included records of the primary search, i.e., all records that meet the inclusion criteria of the review after full-text screening of the primary search results. However, there may be situations in which it is justified to deviate from this recommendation and either use further pertinent records as additional seed references or only a defined sample of the included records.**

Rationale and explanatory details:

The more seed references are used, the better are the chances that citation tracking finds additional relevant unique records. While using only a sample of the included records as seed references may be enough, there is currently no evidence that could help decide how many seeds are needed or how to decide which may perform better. Hence, we recommend using all the records that meet the inclusion criteria of the review after full-text screening of the primary search results.

However, review authors may deviate from this recommendation if they deal with a very small or large number of included records. A very small number of included records may not yield additional relevant records or only have limited value. In this case, review authors could use further records for citation tracking (e.g., systematic reviews on the topic that were flagged during the screening phase). A very large number of included records would lead to too many records to screen. In this case, review authors could use a selected sample of included records for citation tracking (e.g., using an appropriate sampling method). In case of such deviation, authors should describe their rationale and sampling method (e.g., random sample).

Recommendation 4 (Not included in Delphi round 3)

**Backward citation tracking should ideally be conducted by screening the titles and abstracts of the seed references as provided by a citation index. Screening titles as provided when checking reference lists of the seed references can still be performed.**

Rationale and explanatory details:

Citation tracking workflows encompass two consecutive steps: retrieval of records and screening of retrieved records for eligibility. When using an electronic citation index for citation tracking, retrieval and screening are usually separated. While forward citation tracking requires a citation index, backward citation tracking can also be performed by manually checking the reference lists of the seed references. Manual backward citation tracking is sometimes part of an established workflow, e.g., done during eligibility-checking of the full-text record or during data extraction. Merging these two steps has the benefit that researchers know the context in which a reference was used and that all references can be screened. However, manual backward citation tracking has three disadvantages: (i) The retrieval and screening phases are no longer separated which makes reporting of the methods/results difficult and unclear, (ii) the citation tracking results cannot be deduplicated against each other and/or against the primary search results which may add an unnecessarily high workload (see recommendation 6), and (iii) the eligibility assessments are restricted to the titles (instead of titles and abstracts) which could lead to relevant records being overlooked due to unspecific titles mentioned in vague contexts.

In recent years, online citation tracking options via citation indexes or free to access citation tracking tools have become more readily available leading to faster and easier procedures. More and even better tools to facilitate this workflow are expected in the future. Combining citation tracking via citation indexes with automated deduplication (free online tools available) makes this recommendation feasible. A caveat is that a search in a single citation index will in most cases fail to retrieve all the cited references. Thus, references to some documents (such as websites, registry entries or grey literature) that are less likely to be indexed in databases may only be retrievable by checking reference lists or only in some citation indexes.

Recommendation 5

**Using the complementary coverage of two citation indexes for citation tracking to achieve more comprehensive coverage should be considered if access is available. This is especially meaningful if seed references cannot be found in one index and reference lists were not checked.**

Rationale and explanatory details:

A single citation index may not cover all seed references and it very likely will not find all the citing and cited literature. The reason for that is that citation indexes do not offer 100% coverage as some references are currently not indexed in one or several citation index(es). Evidence indicates that when using more than one citation index for citation tracking, the results of the different indexes complement each other. Thus, retrieval of backward and forward citation tracking results from more than one citation index (e.g., TheLens via citationchaser, Scopus, citation indexes in Web of Science) followed by deduplication (see recommendation 6) is a powerful method to increase the sensitivity of citation tracking. It is similar to the complementary effect of using multiple electronic databases for the primary database search, which is the gold-standard in systematic search workflows. In recent years, online citation tracking options have increased and many open access tools make rapid electronic citation tracking universally accessible.

Recommendation 6 (Not included in Delphi round 3)

**Before screening, the results of supplementary backward and forward citation tracking should be deduplicated.**

Rationale and explanatory details:

The concept of citation tracking as a supplementary search method relies on the notion that reference and cited-by lists of eligible references are topically related to these references. This implies a considerable degree of overlap within these lists leading to several duplicates. Furthermore, the overlap likely also extends to the results of the primary systematic search that was performed on the same topic. Based on these considerations and on the fact that the results of the primary search have already been screened for eligibility, the screening load of citation tracking results can be significantly cut by removing those references that have already been screened for eligibility (deduplication against the primary search) and those that appear as duplicates during citation tracking. Depending on the method of deduplication, this can be done in one go.

While deduplication can be conducted manually, nowadays standard bibliographic management software and specialized tools provide automated deduplication solutions, allowing for easier and faster processing.

If citation tracking leads to only a very small number of results, omission of the deduplication step can be considered to save time and administrative effort.

Recommendation 7

**If citation tracking finds additional eligible records, another iteration of citation tracking should be considered using these records as new seed references.**

Rationale and explanatory details:

Citation tracking methods can be conducted over one or more iteration(s), a process we refer to as iterative citation tracking. The first iteration is based on the original seed references (see recommendation 3). If eligibility screening of the results of this first iteration leads to the inclusion of further eligible records, these records serve as new seed references for the second iteration and so forth. There is evidence that conducting iterative citation tracking can contribute to the identification of more eligible records.

Since iterations beyond the first round of citation tracking require additional time and effort and may interrupt the ongoing review process, the decision in favor of or against further iterations should be guided by a cost-benefit assessment. Relevant factors to be assessed include the review topic (difficult-or easier-to-search-for), sensitivity of the primary search, aim for completeness of the literature search, and the estimated potential benefit of the iteration(s) (e.g., based on the number/percentage of included records found with the previous citation tracking iteration).

Review authors are encouraged to report the number of iterations and possibly the reason for stopping if the last iteration still retrieved additional eligible records.

Please note that stating “citation tracking was done on all included records” can lead to confusion. Most authors may mean all records included after full-text screening of the primary search results. However, strictly speaking, “all included records” also includes the records retrieved via citation tracking. The latter interpretation implies that iterative citation tracking is required until the last iteration leads to no further identification of eligible records.

As outlined in the rationale of recommendation 6, results of citation tracking iterations can be deduplicated against all previously retrieved records to reduce the screening load.

Recommendation 8

**Stand-alone citation tracking should not be used for literature searches that aim at completeness of recall.**

Rationale and explanatory details:

We refer to stand-alone citation tracking when any form of citation tracking is used as the primary search method without comprehensive prior database searching. This is contrary to citation tracking as a supplementary search method to a primary database search. Seed references for stand-alone citation tracking could, for example, be records from researchers’ personal collections or retrieved from less sensitive literature searches. Stand-alone citation tracking can be based on a broad set of seed references. It can comprise backward and forward citation tracking as well as indirect methods that collect co-citing and co-cited references.

When study authors replicated published systematic reviews with stand-alone citation tracking, they mostly missed literature that was included in the systematic review. Since search methods for systematic reviews and scoping reviews should aim at completeness of recall, stand-alone citation tracking is not a suitable method for these types of literature review.

Recommendation 9

**Reporting of citation tracking should clearly state**

-**the seed references (if they differ from the set of included records from the results of the primary search along with a justification for this difference),**

-**the directionality of searching (backward, forward, co-cited, co-citing),**

-**the date(s) of searching (which may differ between rounds of iterative citation tracking) (not applicable for reference list checking),**

-**the number of citation tracking iterations,**

-**all citation indexes searched (e.g.,** LENS.ORG**, Google Scholar, Scopus, citation indexes in Web of Science) and, if applicable, the tools that were used to access them (e.g., Publish or Perish, citationchaser),**

-**if applicable, information about the deduplication process (e.g., manual/automated, the software or tool used),**

-**the method of screening (i.e. statement whether the records were screened in the same way as the primary search results or description of the alternative method used), and**

-**the number of citation tracking results in the right column box of the PRISMA 2020 flow diagram.**

Rationale and explanatory details:

The relevant guidance for researchers conducting citation tracking in systematic literature searching can be found in item 5 “citation searching” of PRISMA-S (https://doi.org/10.5195/jmla.2021.962). Accordingly, required reporting items are the directionality of citation tracking (examination of cited or citing references), methods and resources used for citation tracking (bibliographies in full text articles or citation indexes), and the seed references that citation tracking was performed upon. Additional information for the reporting of citation tracking can be found in items 1 (database name), 13 (dates of searches), and 16 (deduplication). While PRISMA-S can be seen as minimum reporting standard for citation tracking as supplementary search technique, other important elements that emerged from our scoping review (https://doi.org/10.1101/2022.09.29.22280494) need to be reported to achieve full transparency and/or reproducibility. These elements are listed in recommendation 9 as a supplement to PRISMA-S to comprehensively guide the reporting of supplementary citation tracking in systematic literature searching.

Concerning reporting of citation tracking results in the PRISMA 2020 flow diagram, two variants are possible: (i) reporting of deduplicated records only which are additional to the primary search results or (ii) reporting of all retrieved records followed by insertion of an additional box where the number of deduplicated records is reported.

Please note that not all citation tracking reporting items have to be reported in the main methods of a study. Detailed search information can usually be provided in an appendix or an online public data repository.

**Part 3: Amended aspects needing further research**

Aspect 1 (Not included in Delphi round 3)

**The effectiveness, applicability and conduct of indirect citation tracking methods as supplementary search methods in systematic reviewing require further research (including retrieval of additional unique references, their relevance for the review and prioritization of results).**

Rationale: Indirect citation tracking involves the collection and screening for eligibility of records that share references in their bibliography or citations with one of the seed references (i.e., co-citing or co-cited references). Indirect citation tracking typically retrieves a large volume of records to be screened. Therefore, prioritization algorithms for the screening of records and cut-offs that may discriminate between potentially relevant and non-relevant records have been proposed that aim at reducing the workload of eligibility screening. The methodological studies that have pioneered indirect citation tracking methods for health-related topics have so far exclusively focused on stand-alone citation tracking. It is currently unclear if the added workload and resources for searching and screening warrant indirect citation tracking methods as supplementary search techniques in systematic reviews of any type (qualitative/quantitative, difficult/easier-to-search-for).

Aspect 2 (Not included in Delphi round 3)

**Further research is needed to assess the value of citation tracking. Potential research topics could be:**

-**influence of citation tracking on results and conclusions of systematic evidence syntheses,**

-**topics or at least determinants of topics where citation tracking likely/not likely has additional value, or**

-**economic evaluation of citation tracking to assess the cost and time of conducting CT in relation to its benefit.**

Aspect 3 (Not included in Delphi round 3)

**Further research is needed to assess the best way to perform citation tracking. Potential research topics could be:**

-**optimal selection of seed references,**

-**optimal use of indexes and tools and their combination to conduct citation tracking,**

-**methods and tools for deduplication of citation tracking results,**

-**subjective influences on citation tracking (e.g., experience of researcher, prevention of mistakes), or**

-**reproducibility of citation tracking.**

Aspect 4 (Not included in Delphi round 3)

**Further research is needed to reproduce existing studies: Any recommendations in this Delphi that are based on only 1-2 studies require reproduction of these studies in form of larger, prospectively planned studies that grade the evidence for each recommendation and propose additional research where the grade of evidence is weak.**

**Part 4: Other comments**

*Provide any other comment as deemed relevant*.

**DELPHI SURVEY ROUND 4**

We amended the recommendations according to the feedback of the Delphi panel. All previous recommendations are now consented (>75% agreement with no relevant changes necessary) and are not to be rated in Delphi round 4 any longer.

Delphi round 4 comprises two official and one in-official tasks:

1. Terminology: The voting for a terminology set in Delphi round 3 returned the following results: The winner set is set 1 with a total of eleven votes (set 2a zero votes; set 2b two votes; set 3 seven votes). To quantify consensus for this terminology set, we now present a new recommendation 1 and ask you to rate your agreement and critically review the associated “rationale and explanation” text. Please note that the wording of recommendations and explanatory texts 2 – 10 was adapted to terminology set 1.
2. All explanatory texts to the recommendations and aspects needing further research have been subjected to draft referencing and the corresponding list of references appended at the bottom of the text. We ask you to review the referencing and suggest additional references where considered appropriate.

**Part 1: Amended draft recommendations**

Recommendation 1

**The following terminology should be used to describe search methods that exploit citation relationships:**

–”Citation Searching” as umbrella term,
– *”Backward Citation Searching”* to describe the sub-method retrieving and screening cited references,
– *”Reference List Checking”* to describe the sub-method retrieving and screening cited references by manually reviewing reference lists,
– *”Forward Citation Searching”* to describe the sub-method retrieving and screening citing references,
– *”Co-cited Citation Searching”* to describe the sub-method retrieving and screening co-cited references,
– *”Co-citing Citation Searching”* to describe the sub-method retrieving and screening co-citing references,
– *”Iterative Citation Searching”* to describe one or more repetition(s) of a search method that exploits citation relationships, and
– *”Seed References”* to describe relevant articles known beforehand.

Rationale and explanation:

As compiled in a recent scoping review [1], the reporting of citation searching methods is frequently unclear and far from being standardized. For example, “citation searching”, “snowballing”, or “co-citation searching” are sometimes used as methodological umbrella terms but also to denote a specific method such as backward or forward citation searching [1]. For clarity, standardized vocabulary is needed.

The set of terms brought forward in this recommendation is the direct result of a Delphi consensus procedure among an international expert panel of systematic review methodologists and information specialists. The set is consistent in itself as well as with the terminology used in PRISMA-S and PRISMA 2020 guidelines [2, 3] and hence well suited for uniform reporting of citation searching.

Recommendation 2 (Final consent 100% – no further modifications)

**For “difficult-to-search-for” systematic search topics, backward and forward citation searching should be seriously considered as supplementary search techniques.**

Rationale and explanation:

Evidence indicates that the ability of citation searching as a supplementary search technique to find additional unique records in a systematic literature search varies with the topic [1]. Searches for particular study designs (qualitative, mixed-method, observational, prognostic, or diagnostic test studies) or health science topics such as non-pharmacological, non-clinical, public health, policy making, service delivery, or alternative medicine have been linked with effective supplementary citation searching. The underlying reasons are manifold and include poor transferability of the topic to text-based searching (e.g., owing to poor conceptual clarity, inconsistent terminology, or vocabulary overlaps with other topics) [4]. The ability of citation searching to find any publication type including unpublished or grey literature or literature that is not indexed in major databases (e.g., concerning a developing country) may also be relevant [5]. However, a clear categorization of “difficult-to-search-for” topics is currently not possible and it remains for the review authors themselves to judge whether their review topic is likely to fall into this category.

We recommend that persons conducting the search who have difficulty assessing whether the topic is difficult-or easier-to-search-for always opt for citation searching or consult an experienced information specialist [6]. If for whatever reason the search strategy does not comprehensively capture the topic, backward and forward citation searching may compensate the potential loss of information at least to some extent.

*Please add any additional referencing suggestions you might have:*

Recommendation 3 (Final consent 86% – no further modifications)

**For “easier-to-search-for” systematic search topics covered by a highly sensitive search, backward and forward citation searching are not explicitly recommended as supplementary search techniques. Reference list checking of included records can be used to confirm the sensitivity of the search strategy.**

Rationale and explanation:

Evidence indicates that the ability of citation searching as a supplementary search technique to find additional unique references in a systematic literature search varies with the search topic [1]. Searches for clearly defined clinical interventions as part of Participant-Intervention-Comparison-Outcome (PICO)-questions have been linked with less effective supplementary citation searching, especially when the search strategies are sensitive and conducted in several databases. However, a clear categorization of “easier-to-search-for” topics is currently not possible and it remains for the review authors themselves to judge whether their review topic is likely to fall into this category.

By checking reference lists within the full-texts of seed references, review authors can test the sensitivity of their primary search strategy (i.e. electronic database search) [7]. Should no additional relevant, unique studies be found, the primary search may have been sensitive enough. Should additional relevant, unique studies be found, it may be an indication that the primary search was not sensitive enough.

We recommend that persons conducting the search who have difficulty assessing whether the topic is difficult-or easier-to-search-for opt for citation searching or consult an experienced information specialist [6]. If for whatever reason the search strategy does not comprehensively capture the topic, backward and forward citation searching may compensate the potential loss of information at least to some extent.

*Please add any additional referencing suggestions you might have:*

Recommendation 4 (Final consent 90% – no further modifications)

**Backward and forward citation searching as supplementary search techniques should be based on all included records of the primary search, i.e., all records that meet the inclusion criteria of the review after full-text screening of the primary search results. Occasionally, it can be justified to deviate from this recommendation and either use further pertinent records as additional seed references or only a defined sample of the included records.**

Rationale and explanation:

The more seed references are used, the better are the chances that citation searching finds additional relevant unique records. While using only a sample of the included records as seed references may be enough, there is currently no evidence that could help decide how many seeds are needed or how to decide which may perform better. Hence, we recommend using all the records that meet the inclusion criteria of the review after full-text screening of the primary search results.

However, review authors may deviate from this recommendation if they deal with a very small or large number of included records. A very small number of included records may not yield additional relevant records or only have limited value. In this case, review authors could use further records as seed references for citation searching (e.g., systematic reviews on the topic that were flagged during the screening phase) [8]. A very large number of included records could lead to too many records to screen. In this case, review authors may use a selected sample of included records as seed references for citation searching. In case of such deviation, authors should describe their rationale and sampling method (e.g., random sample).

*Please add any additional referencing suggestions you might have:*

Recommendation 5 (Final consent 83% – no further modifications)

**Backward citation searching should ideally be conducted by screening the titles and abstracts of the seed references as provided by a citation index. Screening titles as provided when checking reference lists of the seed references can still be performed.**

Rationale and explanation:

Citation searching workflows encompass two consecutive steps: retrieval of records and screening of retrieved records for eligibility. When using an electronic citation index for citation searching, retrieval and screening are usually separated. While forward citation searching requires a citation index, backward citation searching can also be performed by manually checking the reference lists of the seed references. Reference list checking is sometimes part of an established workflow, e.g., done during eligibility assessment of the full-text record or during data extraction [7]. Merging these two steps has the benefit that researchers know the context in which a reference was used and that all references can be screened. However, reference list checking has three disadvantages: (i) the retrieval and screening phases are no longer separated which makes reporting of the methods/results difficult and unclear, (ii) citations from reference list checking cannot be deduplicated against each other and/or against the primary search results which may add an unnecessarily high workload (see recommendation 7), and (iii) the eligibility assessments are restricted to the titles (instead of titles and abstracts) which could lead to relevant records being overlooked due to unspecific titles mentioned in vague contexts. In recent years, online citation searching options via citation indexes or free to access citation searching tools have become more readily available leading to faster and easier procedures [9-11]. More and even better tools to facilitate this workflow are expected in the future. Combining citation searching via citation indexes with automated deduplication (free online tools available [12-14]) makes this recommendation feasible. A caveat is that a search in a single citation index will in most cases fail to retrieve all the cited references. Thus, references to some documents (such as websites, registry entries or grey literature) that are less likely to be indexed in databases may only be retrievable by checking reference lists or only in some citation indexes [15].

*Please add any additional referencing suggestions you might have:*

Recommendation 6 (Final consent 85% – no further modifications)

**Using the combined coverage of two citation indexes for citation searching to achieve more extensive coverage should be considered if access is available. This is especially meaningful if seed references cannot be found in one index and reference lists were not checked.**

Rationale and explanation:

A single citation index or citation analysis tool may not cover all seed references and it very likely will not find all the citing and cited literature. The reasons for that are that citation indexes do not offer 100% coverage as some references are currently not indexed in one or several citation index(es) [16] as well as data quality issues [17]. Evidence indicates that when using more than one citation index for citation searching, the results of the different indexes can complement each other [18-20]. Thus, retrieval of backward and forward citation searching results from more than one citation index or citation analysis tool (e.g., TheLens via citationchaser, Scopus, citation indexes in Web of Science) followed by deduplication (see recommendation 7) can increase the sensitivity of citation searching. It is similar to the complementary effect of using multiple electronic databases for the primary database search, which is the gold-standard in systematic search workflows [21]. In recent years, online citation searching options have increased and many open access tools make rapid electronic citation searching universally accessible [9-11].

*Please add any additional referencing suggestions you might have:*

Recommendation 7 (Final consent 91% – no further modifications)

**Before screening, the results of supplementary backward and forward citation searching should be deduplicated.**

Rationale and explanation:

The concept of citation searching as a supplementary search method relies on the notion that reference and cited-by lists of eligible references are topically related to these references [1]. This implies a considerable degree of overlap within these lists leading to several duplicates. Furthermore, the overlap likely also extends to the results of the primary systematic search that was performed on the same topic. Based on these considerations and on the fact that the results of the primary search have already been screened for eligibility, the screening load of citation searching results can be significantly cut by removing those references that have already been screened for eligibility (deduplication against the primary search) and those that appear as duplicates during citation searching [22]. Depending on the method of deduplication, this can be done in one go.

While deduplication can be conducted manually, nowadays standard bibliographic management software and specialized tools provide automated deduplication solutions, allowing for easier and faster processing [22, 23].

*Please add any additional referencing suggestions you might have:*

Recommendation 8 (Final consent 86% – no further modifications)

**If citation searching finds additional eligible records, another iteration of citation searching should be considered using these records as new seed references.**

Rationale and explanation:

Citation searching methods can be conducted over one or more iteration(s), a process we refer to as iterative citation searching [24]. The first iteration is based on the original seed references (see recommendation 4). If eligibility screening of the results of this first iteration leads to the inclusion of further eligible records, these records serve as new seed references for the second iteration and so forth. There is evidence that conducting iterative citation searching can contribute to the identification of more eligible records [1, 24-26].

Please note that stating “citation searching was done on all included records” can lead to confusion. Most authors may mean all records included after full-text screening of the primary search results. However, strictly speaking, “all included records” also includes the records retrieved via citation searching. The latter interpretation implies that iterative citation searching is required until the last iteration leads to no further identification of eligible records.

*Please add any additional referencing suggestions you might have:*

Recommendation 9 (Final consent 100% – no further modifications)

**Stand-alone citation searching should not be used for literature searches that aim at completeness of recall.**

Rationale and explanation:

We refer to stand-alone citation searching when any form of citation searching is used as the primary search method without extensive prior database searching [1]. This is contrary to citation searching as a supplementary search method to a primary database search. Seed references for stand-alone citation searching could, for example, be records from researchers’ personal collections or retrieved from less sensitive literature searches. Stand-alone citation searching can be based on a broad set of seed references. It can comprise backward and forward citation searching as well as indirect methods that collect co-citing and co-cited references.

When study authors replicated published systematic reviews with stand-alone citation searching, they mostly missed literature that was included in the systematic review [9, 27-29]. Since search methods for systematic reviews and scoping reviews should aim at completeness of recall, stand-alone citation searching is not a suitable method for these types of literature review.

*Please add any additional referencing suggestions you might have:*

Recommendation 10 (Final consent 100% – no further modifications)

**Reporting of citation searching should clearly state**

-**the seed references (along with a justification should the seed references differ from the set of included records from the results of the primary search),**

-**the directionality of searching (backward, forward, co-cited, co-citing),**

-**the date(s) of searching (which may differ between rounds of iterative citation searching) (not applicable for reference list checking),**

-**the number of citation searching iterations (and possibly the reason for stopping if the last iteration still retrieved additional eligible records),**

-**all citation indexes searched (e.g.,** LENS.ORG**, Google Scholar, Scopus, citation indexes in Web of Science) and, if applicable, the tools that were used to access them (e.g., Publish or Perish, citationchaser),**

-**if applicable, information about the deduplication process (e.g., manual/automated, the software or tool used),**

-**the method of screening (i.e., state whether the records were screened in the same way as the primary search results or, if not, describe the alternative method used), and**

-**the number of citation searching results in the right column box of the PRISMA 2020 flow diagram for new or updated systematic reviews which included searches of databases, registers and other sources.**

Rationale and explanation:

The relevant guidance for researchers conducting citation searching in systematic literature searching can be found in item 5 of PRISMA-S [2]. Accordingly, required reporting items are the directionality of citation searching (examination of cited or citing references), methods and resources used for citation searching (bibliographies in full text articles or citation indexes), and the seed references that citation searching was performed upon [2]. Additional information for the reporting of citation searching can be found in PRISMA-S items 1 (database name), 13 (dates of searches), and 16 (deduplication) [2]. While PRISMA-S can be seen as minimum reporting standard for citation searching as supplementary search technique, other important elements that emerged from our scoping review [1] need to be reported to achieve full transparency and/or reproducibility. These elements are listed in recommendation 10 as a supplement to PRISMA-S to comprehensively guide the reporting of supplementary citation searching in systematic literature searching.

Concerning reporting of citation searching results in the PRISMA 2020 flow diagram [30], two variants are possible: (i) reporting of deduplicated records only which are additional to the primary search results or (ii) reporting of all retrieved records followed by insertion of an additional box where the number of deduplicated records is reported.

Please note that the details of the citation searching methods do not have to be reported in the main methods of a study. Detailed search information can be provided in an appendix or an online public data repository.

Examples:

*”As supplementary search methods, we performed […] direct forward and backward CT of included studies and pertinent review articles that were flagged during the screening of search results (on February 10, 2021). For forward CT, we used Scopus, Web of*

*Science, and Google Scholar. For backward CT, we used Scopus and, if seed references were not indexed in Scopus, we manually extracted the seed references’ reference list. We iteratively repeated forward and backward CT on newly identified eligible references until no further eligible references or pertinent reviews could be identified (three iterations; the last iteration on May 5, 2021).” [1]*

*Please add any additional referencing suggestions you might have including further examples of good reporting of citation searching:*

**Part 2: Aspects needing further research**

Aspect 1 (Final consent 92% – no further modifications)

**The effectiveness, applicability and conduct of indirect citation searching methods as supplementary search methods in systematic reviewing require further research (including retrieval of additional unique references, their relevance for the review and prioritization of results).**

Rationale and explanation:

Indirect citation searching involves the collection and screening for eligibility of records that share references in their bibliography or citations with one of the seed references (i.e., co-citing or co-cited references) [31]. Indirect citation searching typically retrieves a large volume of records to be screened [27, 29]. Therefore, prioritization algorithms for the screening of records and cut-offs that may discriminate between potentially relevant and non-relevant records have been proposed that aim at reducing the workload of eligibility screening [9, 28]. The methodological studies that have pioneered indirect citation searching methods for health-related topics have so far exclusively focused on stand-alone citation searching [1]. It is currently unclear if the added workload and resources for searching and screening warrant indirect citation searching methods as supplementary search techniques in systematic reviews of any type (qualitative/quantitative, difficult/easier-to-search-for).

Aspect 2 (Final consent 100% – no further modifications)

**Further research is needed to assess the value of citation searching. Potential research topics could be:**

-**influence of citation searching on results and conclusions of systematic evidence syntheses,**

-**topics or at least determinants of topics where citation searching likely/not likely has additional value, or**

-**economic evaluation of citation searching to assess the cost and time of conducting CT in relation to its benefit.**

Aspect 3 (Final consent 96% – no further modifications)

**Further research is needed to assess the best way to perform citation searching. Potential research topics could be:**

-**optimal selection of seed references,**

-**optimal use of indexes and tools and their combination to conduct citation searching,**

-**methods and tools for deduplication of citation searching results,**

-**subjective influences on citation searching (e.g., experience of researcher, prevention of mistakes), or**

-**reproducibility of citation searching.**

Aspect 4 (Final consent 86% – no further modifications)

**Further research is needed to reproduce existing studies: Any recommendations in this Delphi that are based on only 1-2 studies require reproduction of these studies in form of larger, prospectively planned studies that grade the evidence for each recommendation and propose additional research where the grade of evidence is weak.**

**References (Delphi round 4)**

**APPENDIX 3. TARCiS statement reporting checklist**

**Checklist for reporting of citation searching**

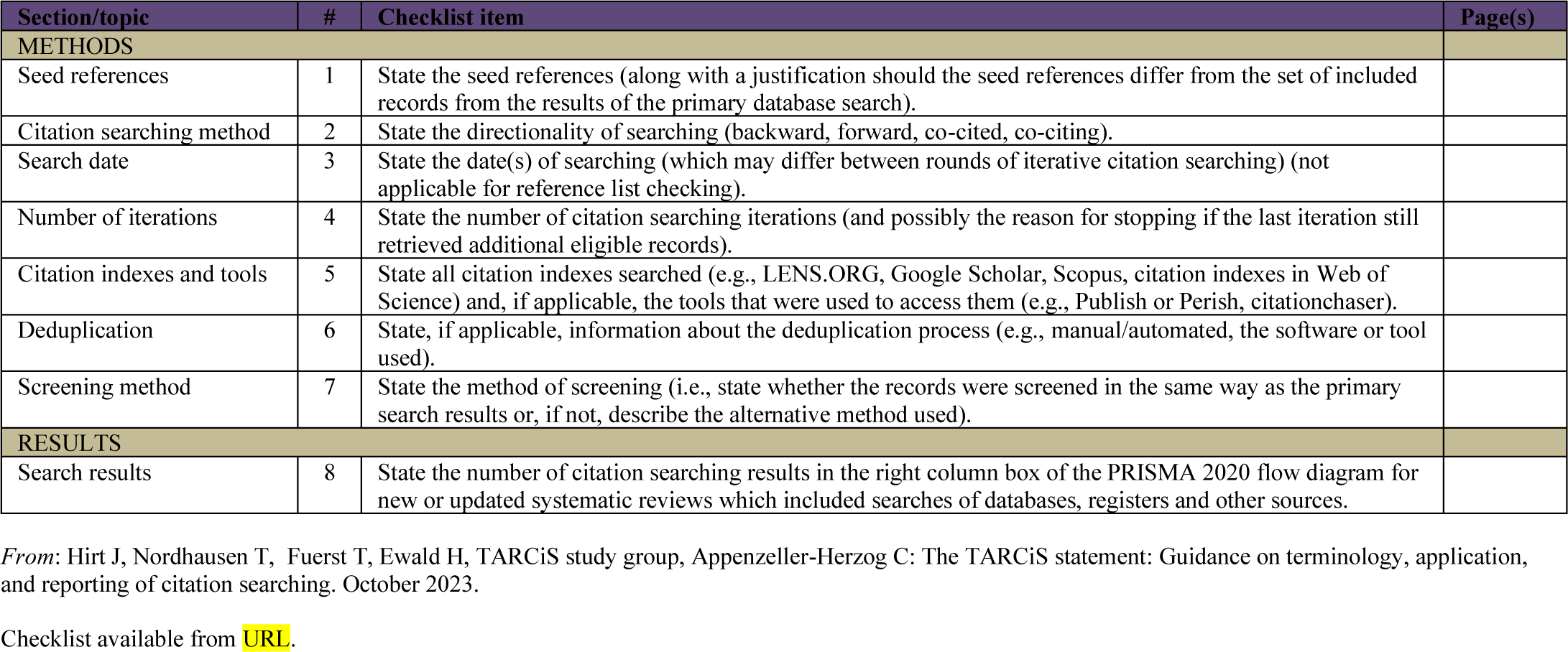

